# Probabilistic Mapping and Automated Segmentation of Human Brainstem White Matter Bundles

**DOI:** 10.1101/2025.05.01.25326687

**Authors:** Mark D. Olchanyi, David R. Schreier, Jian Li, Chiara Maffei, Annabel Sorby-Adams, Hannah C. Kinney, Brian C. Healy, Holly J. Freeman, Jared Shless, Christophe Destrieux, Henry Tregidgo, Juan Eugenio Iglesias, Emery N. Brown, Brian L. Edlow

**Affiliations:** Neuroscience Statistics Research Laboratory, Massachusetts Institute of Technology, Cambridge, MA, USA; Center for Neurotechnology and Neurorecovery, Massachusetts General Hospital, Boston, MA, USA; Institute for Medical Engineering and Science, Massachusetts Institute of Technology, Cambridge, MA, USA; Department of Anesthesia, Critical Care and Pain Medicine, Massachusetts General Hospital and Harvard Medical School, Boston, MA, USA; Athinoula A. Martinos Center for Biomedical Imaging, Massachusetts General Hospital and Harvard Medical School, Charlestown, MA, USA; Department of Neurology, Massachusetts General Hospital, Boston, MA, USA; Department of Pathology, Boston Children’s Hospital and Harvard Medical School, Boston, MA, USA; T.H Chan School of Public Health, Harvard University, Boston, MA, USA; Hawkes Institute, University College London, London, UK; Computer Science and Artificial Intelligence Laboratory, Massachusetts Institute of Technology, Cambridge, MA, USA; Université de Tours, INSERM, Imaging Brain & Neuropsychiatry iBraiN U1253, 37032, Tours, France; CHRU de Tours, 2 Boulevard Tonnellé, Tours, France; The Picower Institute for Learning and Memory, Department of Brain and Cognitive Science, Massachusetts Institute of Technology, Cambridge, MA

**Keywords:** Diffusion MRI, Tractography, Brainstem, Machine Learning, Segmentation

## Abstract

Brainstem white matter bundles are essential conduits for neural signaling involved in modulation of vital functions ranging from homeostasis to human consciousness. Their architecture forms the anatomic basis for brainstem connectomics, subcortical mesoscale circuit models, and deep brain navigation tools. However, their small size and complex morphology compared to cerebral white matter structures makes mapping and segmentation challenging in neuroimaging. This results in a near absence of automated brainstem white matter tracing methods. We leverage diffusion MRI tractography to create BrainStem Bundle Tool (BSBT), which segments eight key white matter bundles in the rostral brainstem. BSBT performs automated segmentation on a custom probabilistic fiber map generated from tractography with a convolutional neural network architecture tailored for detection of small structures. We demonstrate BSBTs robustness across diffusion MRI acquisition protocols through validation on healthy subject *in vivo* scans and *ex vivo* scans of brain specimens with corresponding histology. Using BSBT, we reveal distinct brainstem white matter bundle alterations in Alzheimer’s disease, Parkinson’s disease, and acute traumatic brain injury cohorts through tract-based analysis and classification tasks. Finally, we provide proof-of-principle evidence supporting the prognostic utility of BSBT in a longitudinal analysis of coma recovery. BSBT creates opportunities to automatically map brainstem white matter in large imaging cohorts and investigate its role in a broad spectrum of neurological disorders.

## Introduction

The brainstem is a highly compact structure that orchestrates vital functions such as respiration, circadian rhythm, cardiovascular homeostasis, and consciousness (1–5). These functions are mediated and communicated through the brainstem’s white matter (WM) pathways, whose integrity is increasingly recognized as contributing to both acute neurologic injury and chronic neurodegenerative disease (6–11). Human brainstem connectomics is thus evolving into a thriving topic of research in fields such as network neuroscience, deep brain stimulation and disorders of consciousness (12–16), where a major focus is placed on the accurate delineation of brainstem networks and their connections. With concurrent advances in noninvasive imaging tools such as diffusion MRI (dMRI) tractography (17), brainstem network mapping is now possible to perform *in vivo* and noninvasively. However, because of their small size and complex branching patterns, mapping brainstem WM bundles remains largely unexplored. To address this methodologic barrier and accelerate progress in noninvasive brain connectomics, we aimed to develop a robust, fully automated method to analyze the morphometry and structural integrity of brainstem WM bundles in both healthy brains and those impacted by disease.

Most prior brainstem mapping studies relied upon propagating streamlines between manually labelled brainstem structures with dMRI tractography (14, 16, 18, 19), a time-consuming approach that requires neuroanatomic expertise not widely available. Beyond manual labeling, dMRI segmentation of WM throughout the brain, including certain brainstem WM bundles, is mainly performed through semi-supervised and supervised methods. Semi-supervised approaches include tractography-based aggregation of fibers according to connectivity rules with proximal Regions of Interest (ROIs) (20, 21) (commonly termed virtual dissection), and clustering algorithms that encode feature vectors initialized/modified by user-defined parameters (22, 23). Fully-supervised approaches, which benefit from increased accuracy with sufficient training data, include atlas registration with WM templates, segmentation of automatically generated tractograms constrained by gray matter ROIs, and deep-learning models based on whole-brain tractography (24–27). However, these algorithms have only focused on segmentation of large brainstem WM bundles, such as the corticospinal tracts and superior cerebellar peduncles (25, 27, 28). Most other brainstem WM bundles significantly smaller by volume (19, 29). Furthermore, these bundles generally possess lower dMRI contrast than cerebral hemispheric WM (30) and their contrast is confounded by cardiorespiratory noise and pulsatile flow of cerebrospinal fluid, which creates off-resonance artifacts during *in vivo* MRI acquisition (31, 32) and makes for difficult segmentation. The absence of segmentation tools that focus on smaller brainstem WM bundles thus precludes the automated study of *in vivo* brainstem connectivity in healthy individuals and in patients with neurological diseases that affect the brainstem.

Here, we developed a fully automated, unsupervised brainstem WM bundle segmentation method, which we term *B*rain*S*tem *B*undle *T*ool (BSBT). BSBT segments eight key WM bundles in the pons and midbrain directly from dMRI, without any need for manual intervention.

Specifically, BSBT segmentation is performed on the low-b (b=0 s/mm^2^) and fractional anisotropy (FA) volumes generated from dMRI data, coupled with a multi-channel map generated from automated probabilistic tractography in the brainstem, which is crucial for generating contrast for smaller WM bundles. For segmentation, we utilize a Convolutional Neural Network (CNN) architecture. The CNN possesses two distinct elements that optimize identification of small structures: 1) an attention gate situated between the three highest-resolution encoding and decoding convolutional layers; and 2) a semi-dense conditional random field (CRF) at its *SoftMax* output. We assess BSBT in multiple dMRI domains and varying resolutions through ablation testing in one *ex vivo* and two *in vivo* dMRI datasets, as well as *in vivo* dMRI test-retest analysis in subjects from two scanning sessions. To demonstrate the potential for BSBT to be applied in clinical research and practice, we tested BSBT performance in identifying changes in the intrinsic diffusion characteristics and volume of brainstem WM bundles in three patient cohorts with neurological disorders: Alzheimer’s disease (AD), Parkinson’s disease (PD), and traumatic brain injury (TBI). We then provide proof-of-principle evidence that BSBT yields insights that have the potential to improve prognostication on an individual patient level through longitudinal WM mapping in a severe TBI patient. Finally, we discuss potential applications of BSBT to the study of human brainstem connectivity and the discovery of morphological biomarkers for neurological disorders with brainstem pathology.

## Results

### White matter bundle selection with automated tractographic mapping

We located brainstem WM bundles to segment with BSBT that displayed Probabilistic Fiber Map (PFM) contrast boundaries in both *in vivo* and *ex vivo* dMRI. Details on PFM construction can be found in the methods section and supplementary text. We visually confirmed the presence and identity of each WM bundle with the aid of corresponding Hematoxylin and Eosin/Luxol Fast-Blue (HE-LFB) histological sections in two *ex vivo* brain specimens. We identified the pontine (caudal, c) and mesencephalic (rostral, r) divisions of the medial lemniscus (MLc and MLr), the superior cerebellar peduncle (SCP), the brainstem-based division of the lateral forebrain bundle (LFB), the mesencephalic homeostatic bundle (MHB), the brachium of the inferior colliculus (Bic), and the central tegmental tract (CTG) for BSBT segmentation. In *ex vivo* brain specimens with available histology, we observed PFM contrast boundaries which corresponded with contrast in high-resolution structural MRI and well as in corresponding histological sections (as evidenced by dense, striated, Luxol-Fast-Blue positive regions) for each of the aforementioned WM bundles (Figure 1). In *ex vivo* brain specimens without corresponding histological sections, we relied on corresponding Fast Low-Angle SHot (FLASH) sequences, which we empirically found to contain sufficient contrast to delineate WM bundle boundaries with similar fidelity as histological sections, as illustrated in Figure 1D. PFM intensities in general displayed more reliable and coherent contrast of small, clustered WM bundles in the rostral brainstem than other diffusion maps such as V1, as illustrated in Figure S1. We confirmed the identity of each WM bundle, except for the LFB and MHB, in the *ex vivo* specimens with histology with the Paxinos atlas of the human brainstem (29). Further information on the exact neuroanatomic borders in atlas and FLASH space for each brainstem WM bundle can be found in the supplementary text. Of note, due to the lack of atlas annotations of the LFB and MHB, we assessed their morphology with deterministic tractography, as illustrated in Figures S2-S3.

**Figure 1.**
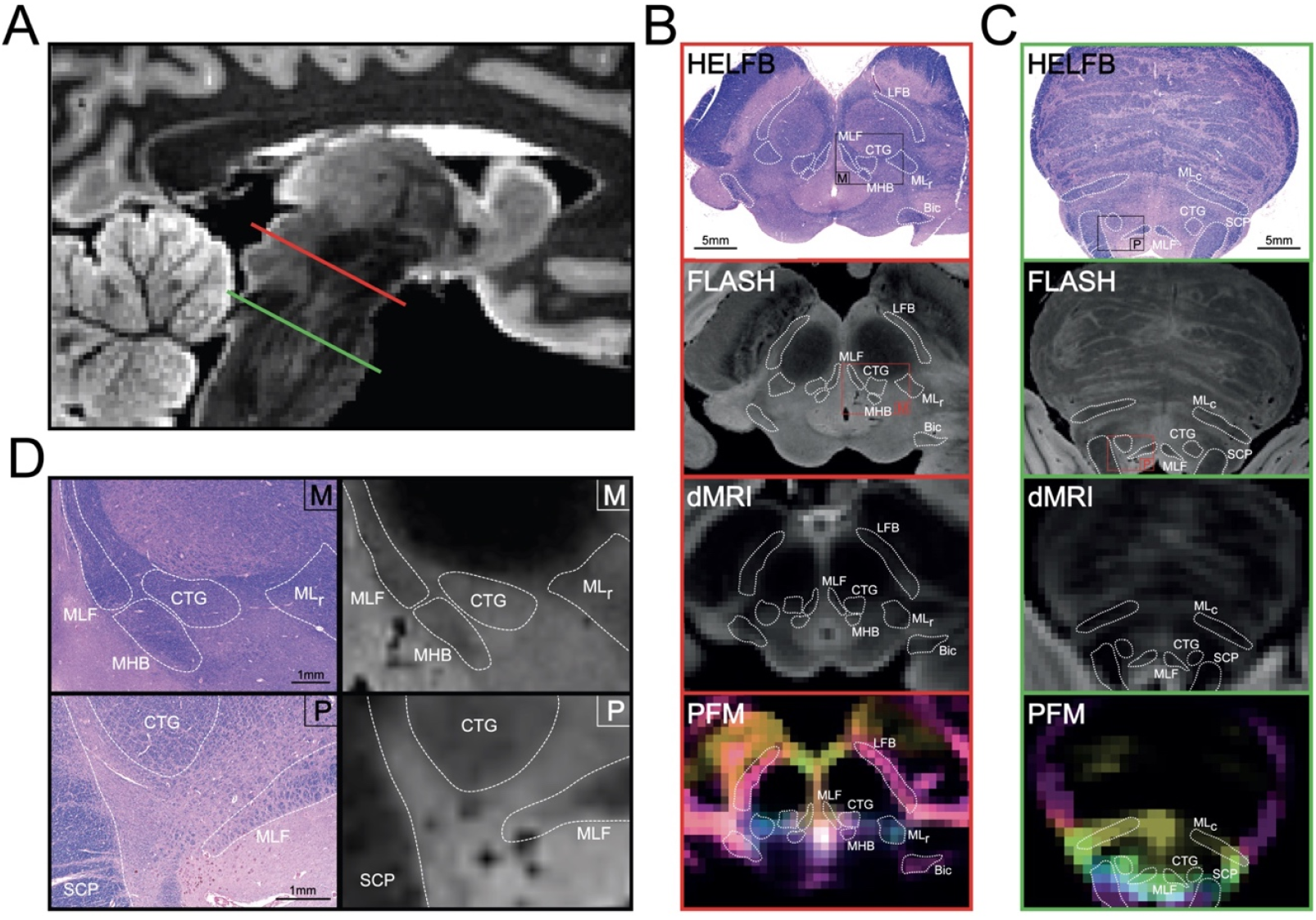
Histological mapping of white matter in the rostral brainstem and correlations with ex vivo MRI and Probabilistic Fiber Mapping. **(A)** A mid-sagittal view of a low-b map from an *ex vivo* brain specimen is shown, with a representative HE-LFB-stained histological section cut along the axial plane of the midbrain at the level red nuclei as indicated by the red slice **(B)**. Shown in **(C)** is a representative HE-LFB histological section at the level of the pons ∼1mm rostral to the roof of the fourth ventricle as indicated by the green slice. For each section, corresponding locations in the FLASH volume, low-b map, and PFM reconstruction are also shown, with the locations of each BSBT WM bundle outlined in white. Zoomed-in views of the midbrain (“M”) and pontine (“P”) histological sections and corresponding FLASH volume contrasts are shown in **(D)**

### Segmentation accuracy compared to ground-truth manual annotations

To test the accuracy of BSBT and its generalizability across different resolutions, scanners, and modalities, we evaluated segmentations on ground-truth brainstem WM bundles that were manually annotated in healthy *in vivo* subjects (n = 20) and normative *ex vivo* brain specimens (n = 7). The annotation process for all specimens is detailed in the supplementary text. Healthy subjects were obtained from the publicly available Human Connectome Project (HCP) WU-Minn 1200 subject release dataset (n = 10, 1.2 mm isotropic resolution, multi-shell dMRI) (33, 34) and from the control group of the third-published Alzheimer’s Disease Neuroimaging Initiative 3 (ADNI3) control dataset (n = 10, 2 mm isotropic resolution, single-shell dMRI) (35). *Ex vivo* brains (750 μm isotropic resolution, single-shell dMRI) were donated by patients without prior neurological disorders who died of non-neurological causes. Clinical and demographic information for each *ex vivo* brain specimen is provided in Table S1. We used Dice scores and average Hausdorff distances (HD) as accuracy metrics for comparison of BSBT segmentations with ground-truth annotations, which are displayed in Figure 2A. A formal description of each accuracy metric can be found in the supplementary text. WM bundles that were larger by volume (SCP, LFB, and Bic) in general displayed higher Dice scores compared to the smaller WM bundles across all three datasets. We observed that HDs were significantly less variable than Dice scores, and these variations were less pronounced with respect to WM bundle size. This is likely attributable to HD being a boundary proximity metric that is less susceptible to fluctuations in overlap, especially between small segmentation labels. The lower HD variability across both scanner resolutions and WM bundle volumes indicates high spatial precision across all resolutions and modalities for BSBT segmentation. Dice scores and HD were better in HCP subjects (mean Dice = 0.70, mean HD = 2.11) compared to ADNI control subjects (subject mean Dice = 0.66, mean HD = 2.34) and *ex vivo* brain specimens (subject mean Dice = 0.62, mean HD = 2.32). Finally, both Dice scores and HD showed relative stability in accuracy until resolutions of 2.5-3mm when synthetically downsampling all datasets, as shown in Figure S4.

**Figure 2.**
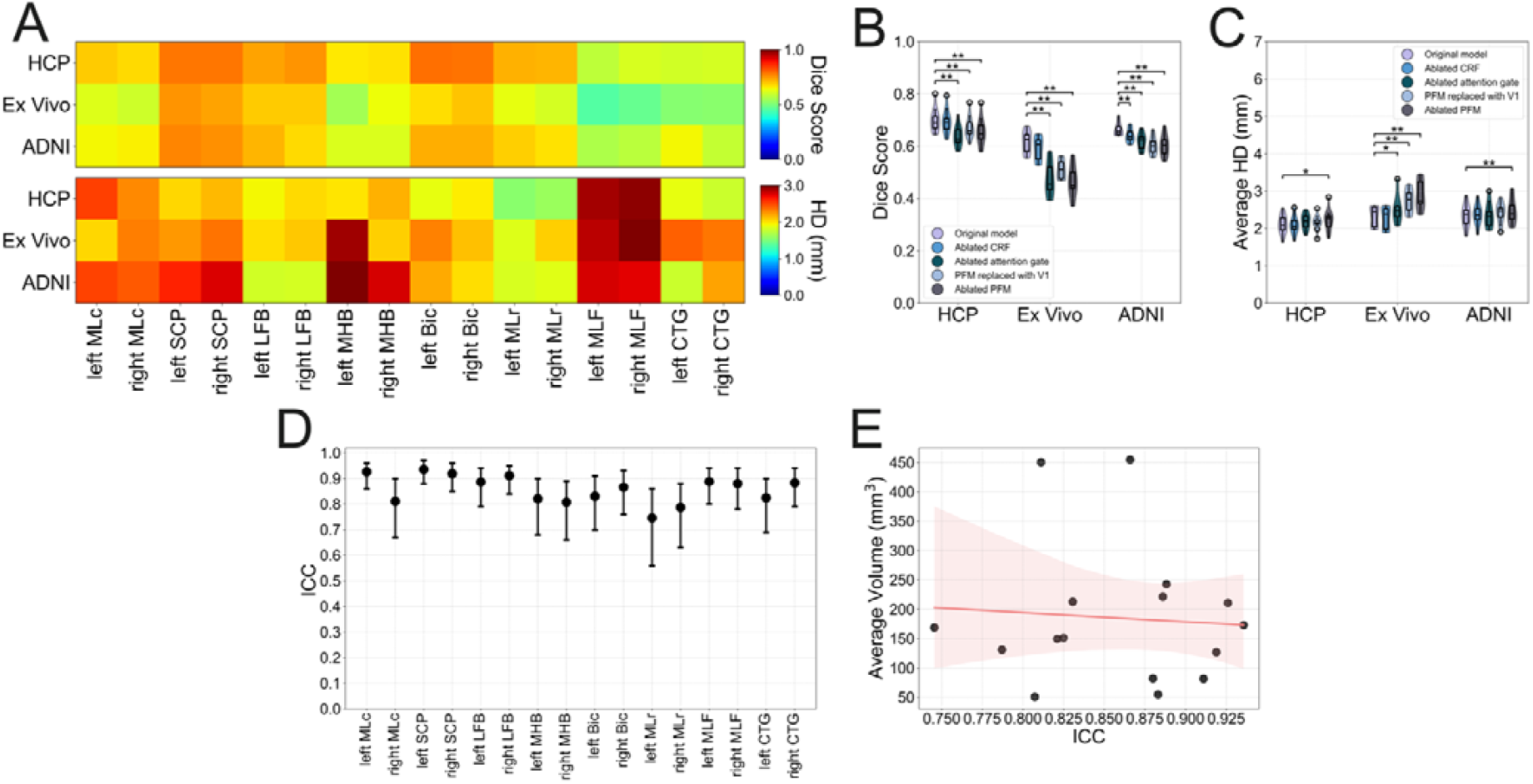
Accuracy, ablation and test-retest reliability analysis for BSBT segmentations. (**A**) Dice scores (top) and HD (bottom) for each brainstem WM bundle CNN segmentation compared to manual annotations for 10 control subjects from the HCP dataset, 7 *ex vivo* brain specimens, and 10 control subjects from the ADNI3 dataset. Each Dice score/HD heatmap element represents a Dice score/HD averaged across subjects within the respective dataset. (**B**,**C**) Violin plots with overlayed box plots consisting of per-subject Dice scores and HD (averaged across all brainstem WM bundles), providing a between-subject comparison of CNN segmentations to manual annotations under varying ablation conditions. Plots are color-coded to represent individual ablations, with significance bars for uncorrected two-tailed Wald tests from a linear mixed effects model: p<0.05 (*) and p<0.01 (**). (**D**) ICC values with 95% confidence intervals, as calculated with a two-way ANOVA mixed effects model, for BSBT-segmented brainstem WM bundle volumes from HCP subjects with test and retest scans. (**E**) ICC values for each brainstem WM bundle plotted against their respective spatial volumes (averaged across all subjects), with a linear regression fit (red line) and shaded 95% confidence interval for the regression line.

### Segmentation accuracy under ablated conditions and test-retest reliability

We performed ablation testing to assess each BSBT component in segmenting brainstem WM bundles. We individually removed the CRF and attention-gating mechanism from the CNN model to test the influence of *SoftMax* correction and attention modulation to localize small, clustered segmentation regions. We removed/replaced the PFM to test the value of vector diffusion maps, such as color FA (V1) and probabilistic tractography, for encoding WM bundle information. Dice scores and HDs for each ablation, along with the unablated scores, are shown in Figure 2 B/C. The unablated CNN resulted in greater segmentation performance (i.e., higher Dice scores and lower HD) than each ablated counterpart. Removal of the attention gating mechanism consistently decreased segmentation performance, with an average Dice score reduction of 0.08 and a 0.1 mm increase in HD across all datasets. This highlights the importance of attention gating for localizing WM structures due to their variable size and sparseness. However, PFM channel removal resulted in the most significant decrease in segmentation performance, with an average Dice score reduction of 0.09 and a 0.3 mm increase in HD across all datasets.

Compared to the original CNN model, all uncorrected p-values for PFM ablation from a linear mixed effects model were <0.01, except for HD in the HCP dataset (p = 0.04). Overall, drop-offs in performance were generally more significant in *ex vivo* data; uncorrected p-values for each ablation except for CRF removal (Dice p=0.08, HD p=0.61) were <0.05. This suggests that with poor diffusion contrast (i.e., fixed brain tissue/weak b-values), enhancing sensitivity to WM boundaries with attention gating and tractography channels is critical to segmentation. Details on the linear mixed effects models used for calculation of significance, as well as acquisition schemes for all HCP, *ex vivo* and ADNI3 MRI data can be found in the supplementary text. In addition to the 15 healthy HCP subjects included for ground truth manual annotation, we analyzed a separate group of 40 HCP subjects who underwent two separate dMRI scanning sessions with the same protocol and average time between scans of 4.8 ± 2.1 months. These subjects served as a “test-retest” group, such that changes in the spatial volume of each brainstem WM bundle between scans were analyzed to assess BSBT segmentation reliability. We observed high reliability for most brainstem WM bundles (ICC > 0.8, Figure 2D). The left/right MLr displayed ICCs > 0.7 (ICC: left = 0.75, right = 0.79). ICCs did not correlate with the average volume of brainstem WM bundles (*R*^2^ = 0.005, *p* =0.79) (Figure 2E), suggesting that BSBT reliability remains consistent across brainstem WM bundles of all sizes. Details on the statistical analysis of each accuracy and reliability metric can be found in the supplementary text.

### Evaluation of diffusion and volumetric changes in neurological disorders

To assess the clinical translatability of BSBT, we evaluated the discriminatory capability of BSBT in AD, PD, and acute severe TBI cohorts based on classification of diffusivity and volume changes of brainstem WM bundles. We calculated voxel-wise average FA inside each bundle ROI as well as average spatial volume as metrics for assessing cohort-wise changes in brainstem WM bundles. FA is widely used and is sensitive to a broad array of pathological changes to the local axonal environment and WM microstructure, and volume is commonly used to assess WM morphology both cross-sectionally and longitudinally in disease states (10, 36–41). To evaluate the discriminatory power of BSBT, we constructed Linear Discriminant Analysis classifiers (henceforth referred to as “classifiers”) on FA and volume of each BSBT WM bundle (n=16). We trained each classifier with leave-one-out cross-validation on healthy and pathological scans from the AD, PD and TBI patient groups. To benchmark BSBT-based classification, we compared Areas Under the Curve (AUC) from the Receiver-Operating Characteristic (ROC) curves of the FA and volume classifiers for brainstem WM bundles to companion classifiers of other commonly segmented brain regions. These include the hemispheric gray matter segmented using *SynthSeg* (42) (n=2), a whole-brainstem mask segmented using *SynthSeg* (n=1), and supra-tentorial WM bundles segmented with *TractSeg* (27) (referred to as *TractSeg* WM bundles) (n=15-16). For more rigorous benchmarking, we chose subsets of *TractSeg* WM bundles with known FA and volumetric changes associated with AD (43, 44) and PD progression (10, 39, 45–50) in dMRI literature. For TBI, we chose *TractSeg* WM bundles with the greatest hemorrhagic lesion burden as observed in our dataset. In the following sections, we report findings for each neurological disorder. Details on MRI acquisition schemes, inclusion/exclusion criteria, subject information, and statistical analysis can be found in the supplementary text.

### Alzheimer’s disease

Loss of WM integrity is a hallmark of AD, and while the vast majority of WM bundles affected are supratentorial, similar changes are observed to extend into brainstem regions (11, 51, 52). Given the potential of brainstem WM to serve as a biomarker for AD and MCI, we assessed brainstem WM bundle FA and volumetric alterations in 106 subjects diagnosed with AD or Mild Cognitive Impairment (MCI) and 122 cognitively normal (control) subjects from the ADNI3 dataset, which we illustrate in Figure 3A/B. FA and log-volume measurements for *TractSeg* WM bundles, hemispheric gray matter masks, and the brainstem mask can be found in Figures S5-S7. All brainstem WM bundles except for the left LFB and left MLr showed reduced median volumes in AD/MCI. The MHB displayed significant volume reductions (left p=0.027, right p=0.024). No brainstem WM bundles exhibited significant differences in average FA between AD/MCI and control cohorts. Brainstem WM bundle classification showed moderate discriminatory performance (AUC=0.60 for volume, AUC=0.58 for FA), and marginally outperformed whole brainstem classification for both volume and FA (AUC=0.58 and AUC=0.54, respectively) without reaching statistical significance (p=0.59 and p=0.42, respectively) (Figure 3D). *TractSeg* WM bundle classification for FA displayed the highest discriminatory power AUC=0.69) (Figure 3C).

**Figure 3.**
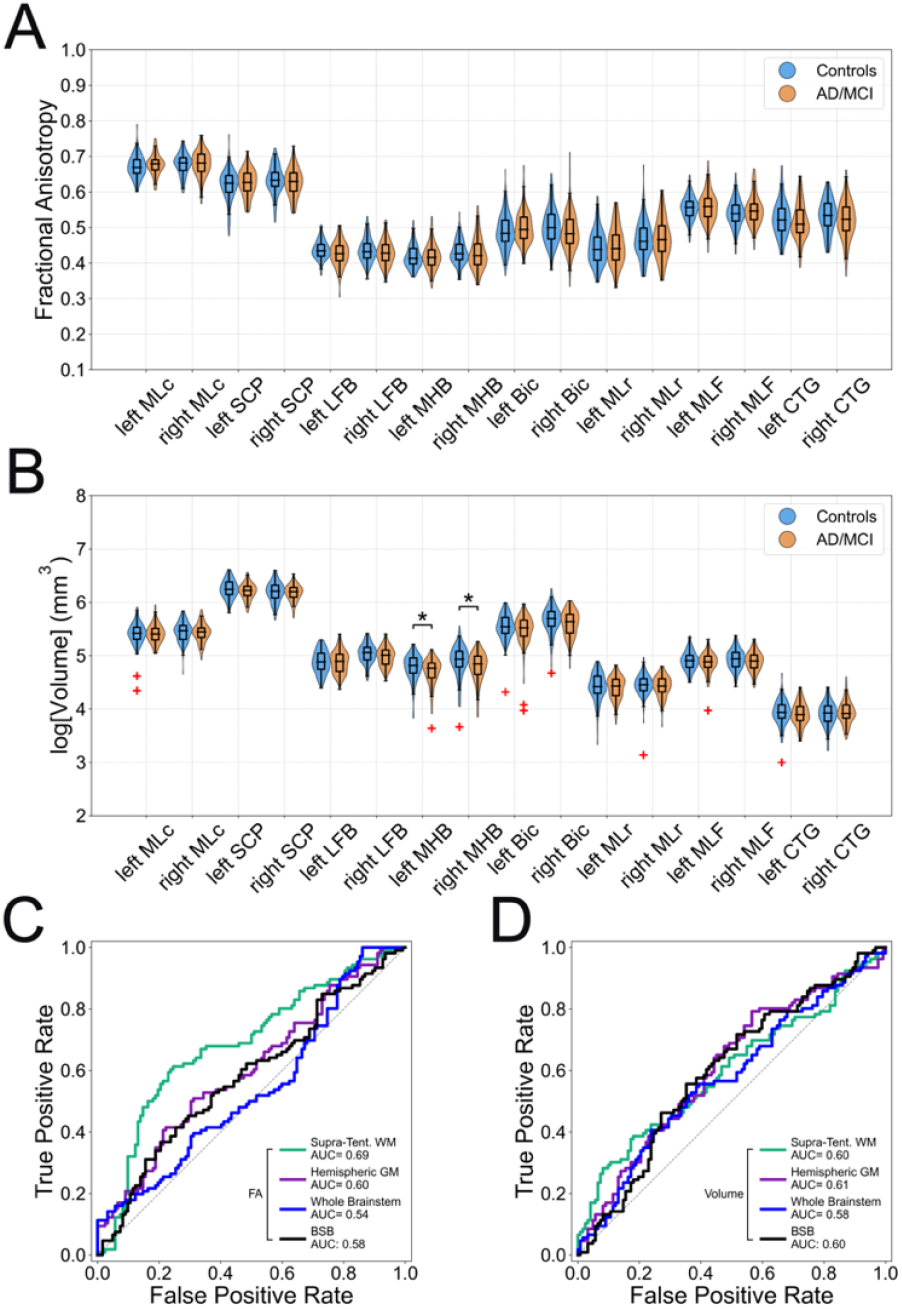
Brainstem white matter bundle alterations and discriminatory analysis in Alzheimer’s disease/mild cognitive impairment. (**A**,**B**) Violin plots of average FA and volume for brainstem WM bundles in the control (blue) and AD/MCI (orange) groups, with significance bars for FDR corrected two-tailed Wilcoxon rank-sum tests: p<0.05 (*) and p<0.01 (**). (**C**,**D**) ROC curves with corresponding AUCs for LDA classifiers trained on FA and volume to distinguish subjects in control versus AD/MCI groups. Classifiers were trained with leave-one-out cross-validation on brainstem WM bundles (n=16) (black), *TractSeg* WM bundles shown to exhibit diffusion/morphological alterations in AD literature (n=15) (green), hemispheric gray matter masks generated with *SynthSeg* (n=2) (purple), and a whole-brainstem mask generated with *SynthSeg* (n=1) (blue). *n* denotes the number of classifier features. GM: gray matter.

### Parkinson’s disease

Many prior studies, including dMRI analyses, have shown PD-related degeneration of brainstem WM. This degeneration is notably encountered in early-stage PD (9, 10, 39, 49, 53) and varies throughout disease progression. We assessed FA and volume changes throughout brainstem WM bundles in 72 patients with PD along with 52 age and sex-matched controls from the Parkinson’s Progression Markers Initiative (PPMI) dataset (54). For the PD patient group, we analyzed dMRI acquired at baseline (at time of diagnosis) and two-year follow-up (2YFU) scanning sessions. FA and log-volume measurements for *TractSeg* WM bundles, hemispheric gray matter masks, and the brainstem mask can be found in Figures S8-S10. BSBT revealed FA reduction between baseline and 2YFU groups in most brainstem WM bundles (Figure 4A). The most significant FA reduction between was observed to be in the LFB (left p<0.001, right p=0.005), Bic (left p<0.001, right p=0.01), and SCP (left p=0.006, right p=0.003). This analysis also revealed bilateral FA reduction in the LFB, Bic and SCP for 75%, 57% and 61% of individual PD subjects between baseline and 2YFU scanning sessions (Figure 4C). Furthermore, FA reduction in the 2YFU group was significant with respect to controls in the left Bic (p=0.006) and left LFB (p=0.046). Volumetric analysis revealed significant volume loss in the MLr (left p=0.002, right p=0.021) between the PD baseline and 2YFU groups (Figure 4B). Furthermore, 63% of the PD individuals showed bilateral volume reduction in the MLr between baseline and 2YFU scanning sessions (Figure 4D). Finally, ROC analysis for three classification scenarios (PD baseline versus control, 2YFU versus control, and PD baseline versus 2YFU) showed that brainstem WM bundle classification either outperformed all classifiers or was comparable to the best classifier in control-2YFU discrimination for FA (AUC=0.69) and volume (AUC=0.63). This performance was statistically significant with respect to the *TractSeg* classifier (AUC=0.53, p=0.024), and approached significance (but not deemed statistically significant) for the whole-brainstem classifier (AUC=0.55, p=0.065) for FA (Figure 4E/F).

**Figure 4.**
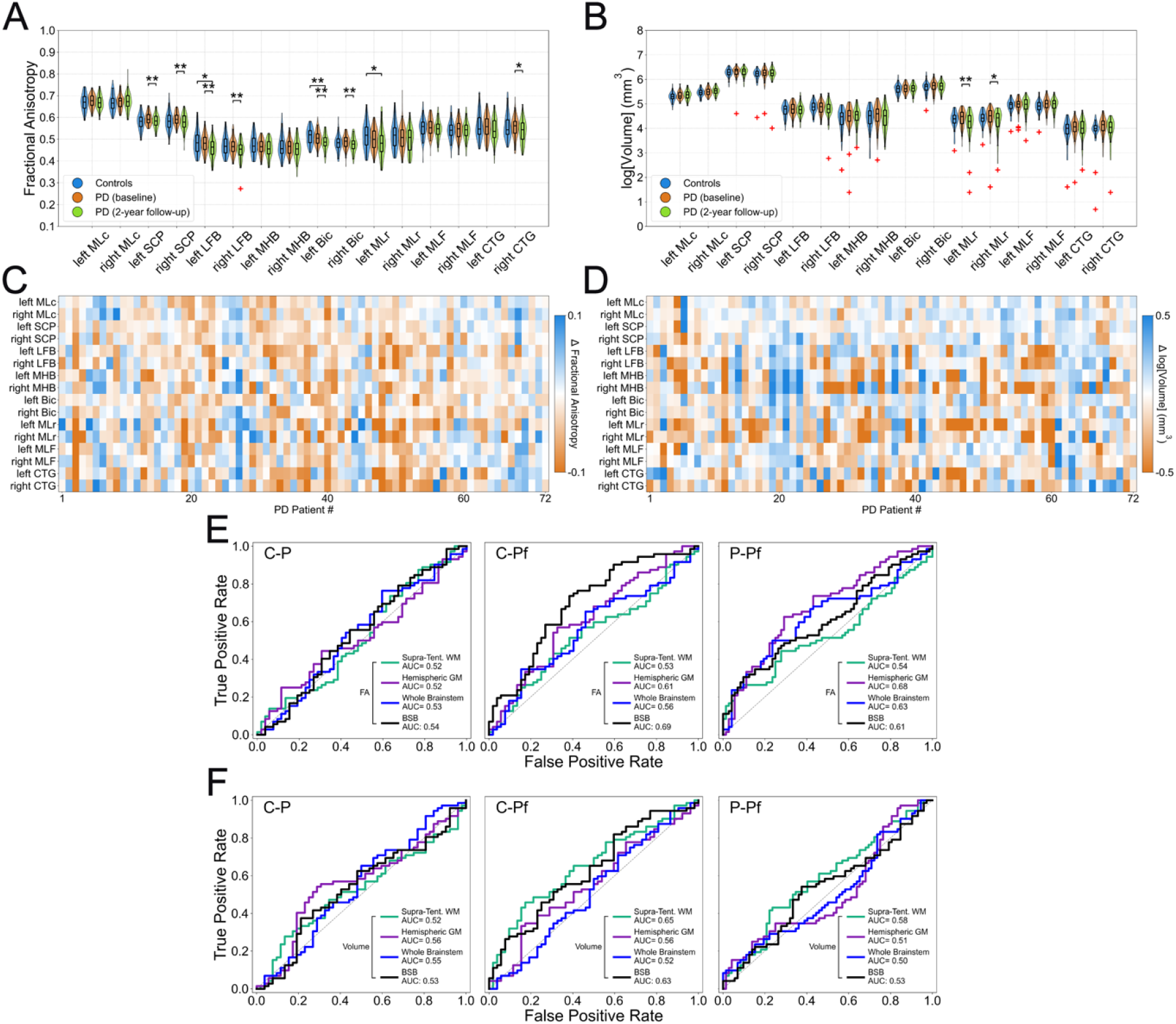
Brainstem white matter alterations and discriminatory analysis in Parkinson’s Disease. (**A**,**B**) Violin plots of average FA and volume for each brainstem WM bundle in control (blue), baseline PD (orange) and 2YFU (green) groups, with significance bars for FDR corrected two-tailed Wilcoxon rank-sum (control-PD and control-2FYU comparisons) and signed-rank (for PD-2YFU comparisons) tests: p<0.05 (*) and p<0.01 (**). (**C**,**D**) Individual changes in average FA and volume for each PD patient between baseline and 2YFU scanning sessions. (**E**,**F**) ROC curves with corresponding AUCs for LDA classifiers trained on FA and volume to distinguish subjects in control versus baseline PD groups (C-P), control versus 2YFU groups (C-Pf), and baseline PD versus 2YFU groups (P-Pf). Classifiers were trained with leave-one-out cross-validation on brainstem WM bundles (n=16) (black), *TractSeg* WM bundles shown to exhibit diffusion/morphological alterations in PD literature (n=15) (green), hemispheric gray matter masks generated with *SynthSeg* (n=2) (purple), and a whole-brainstem mask generated with *SynthSeg* (n=1) (blue). *n* denotes the number of classifier features. GM: gray matter.

### Traumatic brain injury

Severe TBI can lead to multifocal WM disconnection and result in disorders of consciousness, such as coma and vegetative state (55–57). We analyzed FA and volume changes in 17 patients with severe TBI in the acute phase of injury and 29 control subjects to detect early disruption of brainstem WM bundle integrity in disorders of consciousness. Further clinical and demographic information on the TBI patients used for this analysis can be found in Table S2. BSBT analysis revealed no significant changes in volume between TBI and control cohorts while average FA was decreased in the TBI compared to the control group for the majority of brainstem WM bundles (Figure 5A/B). Average FA was significantly lower in three brainstem WM bundles: both LFBs (left p=0.001, right p=0.010), both MLFs (left p=0.033, right p=0.009) and the left MLc (p=0.012). Brainstem WM bundles also showed highly variable overlap with hemorrhagic lesions traced in corresponding susceptibility-weighted images (SWI) in TBI patients, with the CTG displaying the highest overlap score (Dice = 0.113) (Figure 5C). The LFB displayed the greatest lesion overlap score (Dice = 0.062) out of all aforementioned bundles, and the fourth highest overlap out of all brainstem bundles. Details on SWI lesion localization and tract-overlap analysis is described in the supplementary text and illustrated in Figure S11. SWI scans were also used to select *TractSeg* WM bundles for ROC analysis. *TractSeg* WM bundles with the greatest degree of overlap with SWI-traced lesions were used for ROC benchmarking, for which FA and volume measurements are shown in Figure S12. The BSBT classifier for FA outperformed all other FA classifiers, with statistical significance against the hemispheric gray matter classifier (p = 0.035) (Figure 5D). The BSBT classifier for volume outperformed both the hemispheric gray matter and whole-brainstem classifiers, albeit without statistical significance (Figure 5E). FA and log-volume measurements for *TractSeg* WM bundles, hemispheric gray matter masks, and the brainstem mask can be found in Figures S12-S14.

**Figure 5.**
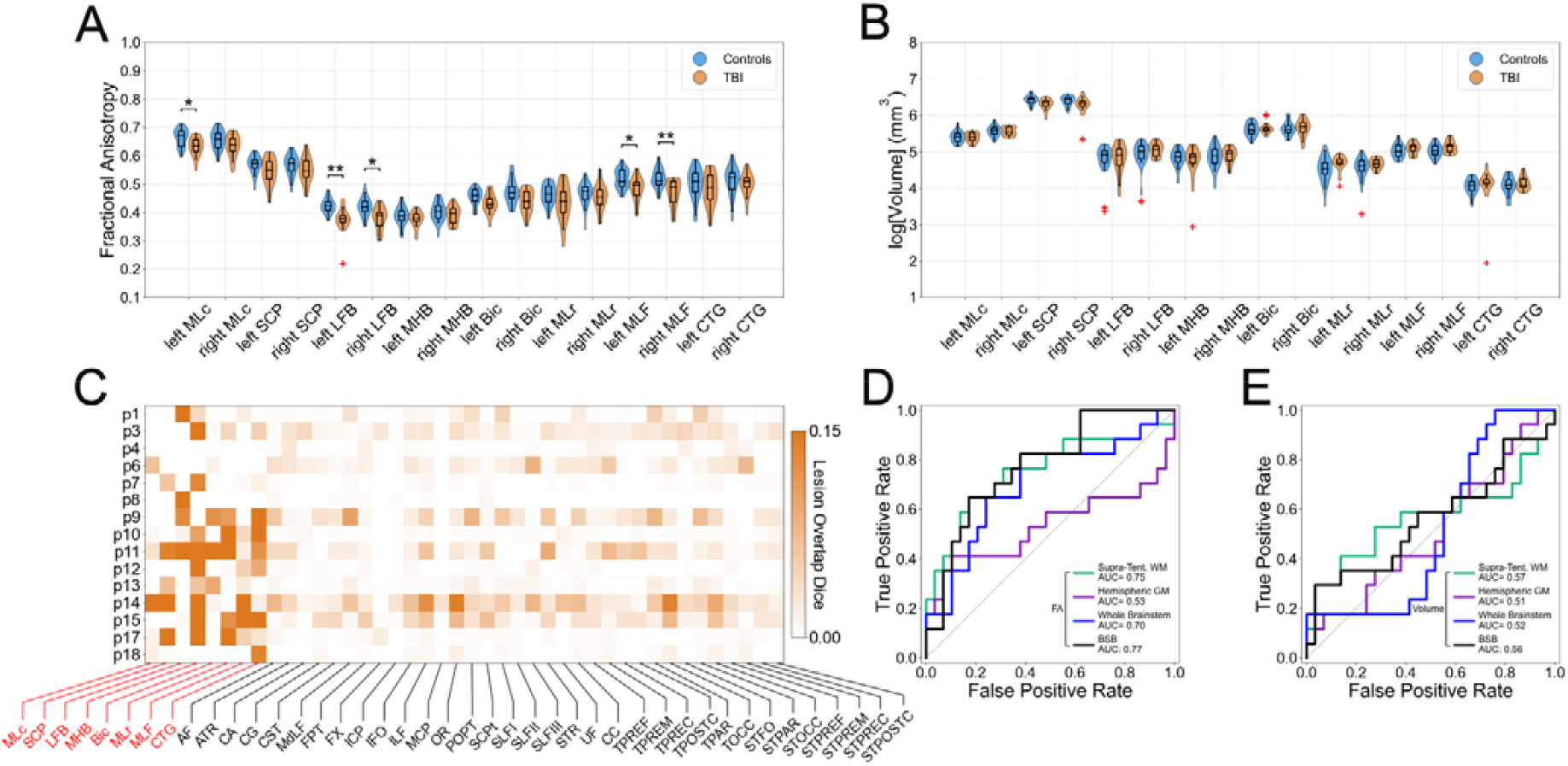
Brainstem white matter alterations, tract-lesion overlap, and discriminatory analysis in Traumatic Brain Injury. (**A**,**B**) Violin plots of average FA and volume for brainstem WM bundles in the control (blue) and TBI (orange) groups, with significance bars for FDR corrected two-tailed Wilcoxon rank-sum tests: p<0.05 (*) and p<0.01 (**). (**C**) Degree of Dice score overlap of WM bundles derived from BSBT and *TractSeg* with hemorrhagic lesions annotated in corresponding SWI volumes. For this comparison we dilated each *TractSeg* ROI with a 2-voxel cube kernel to capture peri-lesional tissue effects. BSBT ROIs were not dilated due to their comparatively small size. (**D**,**E**) ROC curves and corresponding AUCs of LDA classifiers trained on average FA and volume to distinguish control from TBI subjects. Classifiers were trained using leave-one-out cross-validation on brainstem WM bundle (n=16) (black), *TractSeg* WM bundles displaying the greatest degree of overlap with hemorrhagic lesions (n=15) (green), hemispheric gray matter masks generated with *SynthSeg* (n=2) (purple), and a whole-brainstem mask generated with *SynthSeg* (n=1) (blue). *n* denotes the number of classifier features. GM: gray matter.

### Longitudinal white matter bundle morphometry in traumatic coma recovery

We performed a longitudinal analysis of PFM construction and BSBT segmentation in a patient (male in their late 20’s) with an acute disorder of consciousness caused by severe TBI (patient 15 from the TBI dataset, see supplementary Table S2). The patient’s coma was attributed to a large hemorrhagic lesion in the midbrain. Specifically, MRI scanning on day 7 post-TBI revealed an acute traumatic hemorrhage along the entire midsagittal extent of the midbrain (Figure 6A), which typically results in a poor long-term outcome (58). We chose this patient for morphometric analysis for two reasons: First, he had the largest brainstem lesion of any patient in our ongoing study on acute severe TBI (ClincialTrials.gov NCT03504709), and second, he regained consciousness, communication, and partial functional independence by 7 months post-injury (Glasgow Outcome Scale-Extended score of 5) (59). To identify the impact of the midbrain lesion on WM mapping, we segmented the patient’s brainstem WM bundles using the BSBT CNN model with and without a PFM channel. Visual inspection of PFM intensities showed that WM bundles near the lesion periphery were laterally displaced, but not disconnected, by the hemorrhage (Figure 6B). These WM bundles were detected by the CNN model with the PFM, but corresponding segmentation without the PFM channel (i.e., only with low-b and FA inputs) resulted in near-absent labels on the radiologic left side of the midbrain. Thus, the segmentation without the PFM channel indicated severe tract disconnection (Figure 6C). In contrast, the segmentation with the PFM revealed sparing and preservation of brainstem WM bundles providing a potential explanation for the patient’s unexpected long-term functional recovery. A 7-month follow-up MRI revealed a decrease in hemorrhagic lesion volume from 1,962 mm^3^ (acute) to 679 mm^3^ (follow up) (Figure 6D), and the PFM channel showed that displaced WM bundles shifted back towards their expected neuroanatomic locations near the midline (Figure 6E). BSBT CNN models with and without the PFM channels both showed bilateral WM bundle reconstructions at 7-months follow-up (Figure 6F). Collectively, these longitudinal observations imply that the use of standard dMRI channels can lead to false-negative segmentations, whereas PFM information aids in accurate reconstructions of WM bundles in heavily lesioned and deformed brainstem regions.

**Fig. 6.**
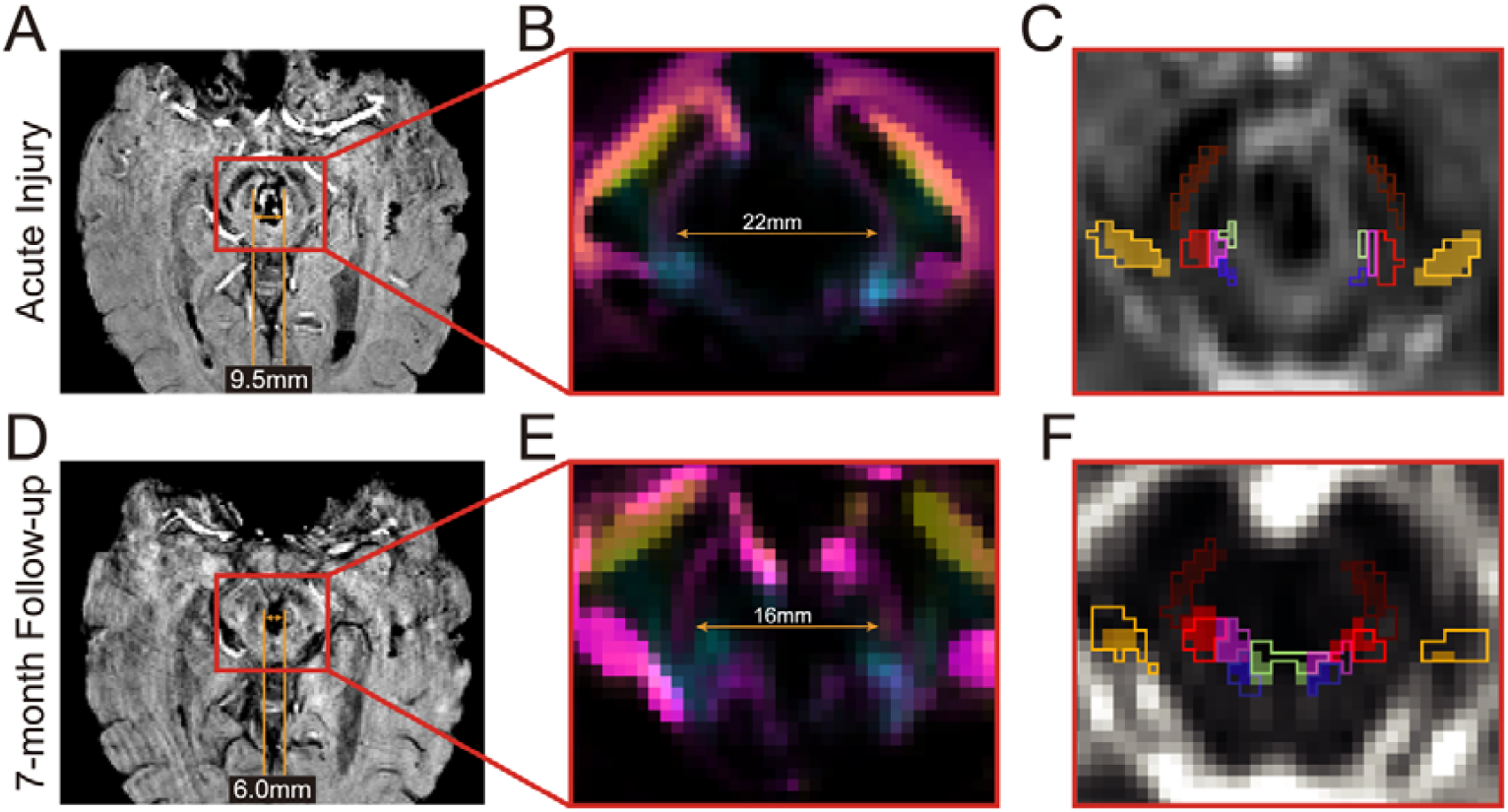
Longitudinal brainstem white matter alterations in a patient with severe traumatic brain injury. **(A)** SWI scan during the acute injury phase of a patient who suffered a traumatic coma from a severe TBI. **(B)** PFM channel from the corresponding dMRI volume, showing significant mass-effect causing displacement, but not direct lesioning, of brainstem WM bundles due to the development of an acute traumatic midbrain hemorrhage. **(C)** The Mean Diffusivity dMRI channel with WM bundle segmentations from a CNN using the PFM channel (outlined), overlayed with semi-transparent segmentations from a CNN without the PFM channel (i.e., segmentation was performed with only the low-b and FA maps). The patient had functional recovery and underwent follow-up scanning at 7 months, which showed a significant decrease in the volume of the midbrain lesion **(D)**. The corresponding PFM channel **(E)** showed a counter-displacement of the brainstem WM bundles proximal to the lesion margin back towards the midline. Due to the undeformed anatomy, BSBT segmentations from a CNN both with and without the PFM channel revealed coherent WM bundle reconstructions **(F)**.

## Discussion

We developed BSBT, an unsupervised algorithm that reliably segments brainstem WM bundles in dMRI scans of the human brain. Through comparison with gold-standard manual annotations in *ex vivo* dMRI, and silver-standard manual annotations in single-and multi-shell *in vivo* dMRI, we found that BSBT segmentations were highly accurate and reliable across multiple dMRI contrasts. We demonstrate the clinical translatability of BSBT and provide proof-of-principle evidence for its use in assessing and classifying neurological conditions that differentially affect the brainstem, such as AD, PD, and TBI. We release BSBT to the research and clinical communities as a tool to map human brainstem WM in healthy and diseased brains (github.com/markolchanyi/BSBT).

BSBT addresses a key gap in the field of brainstem imaging because it is capable of automatically segmenting small brainstem WM bundles, does not require manual intervention, and is generalizable across different dMRI modalities and domains. BSBT showed high segmentation accuracy in both *in vivo* and one *ex vivo* dMRI datasets of varying resolutions (0.7 to 2 mm), with Dice scores ranging from 0.62 to 0.70 and subject-averaged HD not exceeding 2.5 mm. These Dice scores and HDs are comparable to state-of-the art segmentation algorithms of similarly-sized brain regions such as hypothalamic and thalamic nuclei (60, 61). Ablation and test-retest analyses further demonstrated the reliability and consistency of BSBT. Specifically, ablation revealed that coupling unsupervised tractography with a segmentation model modified for enhancement of small regions optimizes segmentation of brainstem WM bundles. This optimization is evidenced through the individual removal of each core BSBT component, both within the CNN model (CRF and attention gating mechanism) as well as the input features (PFM removal/replacement with V1) resulting in a statistically-significant reduction in accuracy in at least one testing dataset. Although segmentation reliability for small regions is expected to be low (due to fluctuations in measured volume from noise), test-retest analyses yielded high intra-subject ICC scores (ICC>0.8) across all brainstem WM bundles, except for the right/left MLC and left MHB (ICC>0.7). This observation implies that BSBT leverages anatomically plausible contrast information that is invariant between scans and not structures that arise from noise. Furthermore, the negligible correlation between ICC and volume of each BSBT-segmented WM bundle suggests that each segmented structure is above the signal-to-noise-ratio limit in HCP-quality data.

The potential clinical utility of BSBT is highlighted by single-tract analysis and multi-tract classification in three disease types: AD, PD, and TBI. More specifically, we explore how acute brain injury and chronically-progressing neurogenerative processes such as in AD and PD exhibit differential effects in brainstem WM integrity both temporally and in terms of underlying diffusion properties. BSBT revealed a unique set of brainstem WM bundles with FA and/or volumetric changes in each disorder. Consequently, diffusion and volume estimates captured through BSBT have the potential to be leveraged as powerful features for classifying neurological disorders with brainstem WM pathology.

In the setting of AD, neurodegeneration initially arises in specific supratentorial nuclei. As the disease progresses, abnormal proteins, such as amyloid-β, are proposed to spread to the brainstem and its associated WM pathways (62, 63). Our BSBT findings provide preliminary evidence for volumetric changes in distal brainstem WM bundles that connect to these supra-tentorial regions. Specifically, BSBT analysis in the ADNI3 AD/MCI cohort revealed volume reduction of the MHB, a bundle connecting several brainstem arousal and homeostatic nuclei, such as the locus coeruleus, to gray matter targets with AD-related degeneration patterns, including the basal forebrain, hippocampus and entorhinal cortex (14, 64–66). We did not identify FA changes in brainstem WM bundles, while supra-tentorial WM displayed more widespread FA/volume reduction (Figure S5) and provided the most powerful LDA classification. These results are unsurprising because brainstem WM changes in AD and MCI is less evident, albeit less-studied, than in supra-tentorial WM (11, 52, 67, 68). However, the observed volumetric changes in the MHB coupled with moderate volume classification performance warrants the use of BSBT to explore other brainstem WM biomarkers, their connectivity to and from brain regions with known AD-related degeneration, their utility in imaging-based AD diagnostics cross-sectionally, as well as longitudinal, dynamic changes as patients transition from prodromal to symptomatic AD.

PD is characterized by accumulation of α-synuclein in the brainstem and ascending, secondary loss of dopaminergic neurons in the substantia nigra among other regions. While brainstem structural degeneration is documented in prior literature (69, 70), the affected WM pathways have not been comprehensively classified. In our longitudinal PD analysis, the most pronounced BSBT finding was bilateral FA reduction in LFB, Bic and SCP. These findings are consistent with and lend support to potential brainstem involvement in the degeneration of nigral, basal ganglia, forebrain, cerebellar and cortical targets associated with these brainstem WM bundles (16, 69, 71– 74). FA patterns followed a bi-phasic trajectory, where FA increased between controls and baseline PD, followed by a decrease in 2YFU PD scans. This longitudinal FA pattern has been previously described in PD dMRI literature (70) and may partially explain the pronounced FA reduction between PD groups relative to the control group. While comparatively fewer WM bundles showed notable FA reduction between controls and 2YFU groups, the left LFB and left Bic showed statistically significant FA decreases in both aforementioned comparisons. The corresponding control-2YFU BSBT classifier also had the highest AUC out of all other classifiers, with an AUC of 0.69. This result lends further support for the involvement of discrete brainstem WM bundles in disease progression and suggests that loss of axonal integrity in discrete brainstem WM bundles, as measured by FA, possess high predictive power as an early-stage PD biomarker. Collectively, these findings highlight how assessment of brainstem WM bundle diffusion metrics can contribute to both PD detection and disease progression.

The application of BSBT to patients with acute severe TBI demonstrates two additional features of the algorithm: its ability to detect focal structural alterations in brainstem WM bundles and to identify bundles in the presence of deformed and/or lesioned brainstems. Pathological and neuroimaging studies suggest that axonal injury in acute severe TBI leads to a decrease in FA with regional variability, and commonly-observed diffusivity changes in proximity to brainstem arousal regions (55, 75–79). In accordance with these findings, BSBT-segmented brainstem WM bundles showed a variable reduction of average FA in the TBI cohort. Importantly, we found statistically significant FA reduction bilaterally in the LFB and MLF and in the left MLc. FA reduction in these bundles likely contributed to the high predictive power of the BSBT classifier for FA, which outperformed every competing classifier in terms of AUC. While there is strong evidence from brainstem lesion studies that injury to arousal nuclei can cause coma (13, 80–82), little is known about which brainstem WM bundles connecting these arousal nuclei to each other and extra-brainstem targets cause disorders of consciousness. The LFB contains connections between brainstem arousal nuclei and parietotemporal regions of the default mode network and is thus believed to be a key pathway that integrates arousal and awareness in human consciousness (16). BSBT identification of LFB FA reduction thus provides direct evidence for WM-centric coma-causing lesions in the brainstem and warrants future investigation into the role of individual brainstem WM pathways in coma pathogenesis.

We also demonstrated that BSBT can identify preserved brainstem WM bundles in a severe TBI patient with a large brainstem lesion who experienced full functional recovery. In the patient’s acute scan, we identified BSBT WM bundles even with high deformation by mass effect – an observation that highlighted the crucial role of the PFM. At the seven-month follow-up dMRI scan, when the lesion size had decreased, BSBT segmentation – both with and without PFM channels as CNN inputs – reconstructed WM bundles on both the left and right margins of the lesion.

Therefore, PFM-based segmentations in the acute scan were anatomically founded and not false-positive reconstructions. We postulate that the high sensitivity of the PFM coupled with a CNN segmentation model has substantial prognostic potential by identifying preserved brainstem bundles that can facilitate coma recovery.

The described evaluation of BSBT and its applications to patients with neurological disorders is limited by model design constraints and the overall composition of the assessed clinical datasets. Our model-specific limitation mainly pertained to the setup of the CNN training protocol, which was limited to 30 training subjects from a single dataset (HCP) annotated by a single rater (MDO) and subsequently posed a risk for model overfitting. To combat overfitting, we aggressively augmented our training data but still observed noticeable accuracy differences between native HCP and *ex vivo*/ADNI test subjects. While this is indicative of model parameters overly-tuned to training data-specific features, ICC between HCP test-retest subjects remained high and segmentation performance for each dataset was consistent across a large resolution span (Figure S4). This result indicates that true anatomic WM boundaries and not features unique to HCP contrast are segmented. Lower dice scores in *ex vivo* test data can potentially be explained by a high *in vivo*-to-*ex vivo* domain shift. *Postmortem* fixation restricts free-water diffusion in brain tissue, which can drastically alter the diffusion signal profile (83) and complicate modelling *ex vivo* contrast from *in vivo* training data. Thus, incorporating other datasets for CNN training, such as *ex vivo*, low-field, and low angular resolution dMRI, would likely increase segmentation robustness.

The primary limitations for all clinical datasets we analyzed were class imbalance and comparatively high feature numbers relative to sample sizes, posing a risk of spurious statistical assessments of WM bundle integrity and classification bias. To combat these confounders, we used non-parametric statistical models with false-discovery rate correction for more rigorous statistical analysis, and implemented a simple, linear model (LDA) for classification to avoid overfitting. However, larger samples sizes for each dataset and classification models with fewer features are still necessary to validate our clinical findings, especially in the setting of multi-bundle analysis. Another limitation of our core analysis which applies to all clinical datasets is the effect of partial voluming on brainstem WM bundles. Given the brainstem’s compactness coupled with coarser *in vivo* scanning resolutions, inclusion of proximal gray matter tissue within each WM bundle ROI is inevitable and likely confounds the identification of pure-WM pathology due to this partial voluming effect. This is particularly pertinent in the presence of gray matter alterations adjacent to the segmented WM bundles, such as PD-related α-synuclein inclusions in the brainstem tegmentum and substantia nigra (which directly borders the LFB) (9, 84), or direct injury to brainstem arousal nuclei such as the ventral tegmental area (adjacent to the LFB/MHB/MLr), periaqueductal gray (adjacent to the MLF/MHB), locus coeruleus (adjacent to the MLF/CTG), or parabrachial complex (adjacent to the SCP/CTG) in traumatic coma (55, 81, 82). The disease-related alterations presented for each brainstem WM bundle therefore need to be interpreted with caution, and future validation studies are needed to determine the spatial limitations WM bundle localization in clinical-grade dMRI. Finally, we note several limitations that were dataset-specific. For PD subjects, relatively low signal-to-noise ratios likely influenced both the number of excluded subjects as well as statistical analysis, as outlined in the methods section and Figure S15. We applied Gaussian smoothing to FA maps, which are in general more noise-susceptible (85), to attempt to match signal-to-noise ratios in the ADNI and TBI datasets. However, spurious statistical changes may have still arisen due to noisy FA measurements, which highlights the need for more robust evaluation with alternative PD dMRI acquisitions and denoising methods.

AD and TBI analyses were likely hindered by the variable distribution of disease stage. To increase sample size, we grouped AD and MCI subjects to an “AD/MCI” supergroup, albeit at the potential expense of capturing longitudinal variability in FA and volume measurements (52, 86).

Similarly, for TBI subjects, the principal longitudinal limitation was variable injury-to-imaging intervals. Because dynamic microstructural alterations occur within hours-to-days of a TBI, the time from injury to imaging becomes a critical factor for diffusion measurements (87). In future work, we aim to standardize TBI patient groups with fixed injury-to-imaging intervals, which would also provide deeper insights into the longitudinal effects of WM injury on brainstem dysfunction and recovery as well as the differential effects of both primary and secondary injury mechanisms.

In summary, we present BSBT, a brainstem WM bundle segmentation algorithm which we rigorously validated with high-resolution *ex vivo* and *in vivo* dMRI. With a CNN heavily modified for small-structures, we provide automated segmentations of key WM bundles in the pons and midbrain which are central to many of the vital functions of the brainstem. We demonstrate the potential of BSBT for classification of neurological disorders (AD, PD, and TBI) and prognostication through assessing brainstem WM morphology in a single dMRI scan. This automated approach to studying brainstem WM bundles stands to streamline neuroimaging research and promote clinical investigation of brainstem WM morphology, integrity, and connectivity in healthy individuals and in neurological disease. While this method is currently focused on eight brainstem WM bundles, there remain many avenues for extending the segmentation model to additional bundles in a straightforward fashion, which we hope will become a backbone for brainstem diffusion analysis and connectivity mapping.

## Materials and Methods

### Probabilistic Fiber Map construction

The workflow to generate the PFM, as well as accompanying deterministic tractography reconstructions for all brainstem WM bundles segmented with BSBT are illustrated in Figure 7. We enable users to only input a dMRI volume without requiring companion structural volumes such as T1 sequence, which is common in dMRI segmentation algorithms (25, 61), by performing all processing in dMRI space. We segment the low-b volume with the FreeSurfer tool *SynthSeg* (42) to generate ROIs of the ventral diencephalon, thalamus, and cerebellar WM, which are used as seed and target regions for the PFM. We then generate a synthetic MP-RAGE from the low-b volume using *SynthSR* (88) to extract the pons and medulla subfields using FreeSurfer’s brainstem subregion segmentation tool (28), which are used as inclusion/exclusion regions for the PFM. Only the infero-lateral margins of the ventral diencephalon ROI are used for PFM construction to minimize overlap with probabilistic streamlines generated from the thalamic mask. This margin is approximated by the intersection of a dilated ventral diencephalon mask and a dilated mask of the *SynthSeg*-segmented amygdala. We perform local probabilistic tractography between three pairs of the *SynthSeg*-generated ROIs to obtain brainstem streamlines to generate the PFM. Streamlines between thalamic and medullary masks form contrast for the first PFM channel. Streamlines between cerebellar WM and ventral diencephalon masks, with a pontine mask is used as an exclusion region, form contrast for the second PFM channel. Streamlines between ventral diencephalon and medullary masks, with a pontine inclusion region cerebellar WM exclusion region (to avoid streamlines entering the medulla through the middle cerebellar peduncle), form contrast for the third PFM channel. 10^5^ tracts are uniformly seeded and propagated with a spline angle of 55° for contrast generation in each PFM channel. Intensity distributions corresponding to streamline density per-voxel from the first two PFM channels are binned and their histograms are linearly matched to the third PFM channel histogram. All dMRI preprocessing steps including denoising, distortion correction and tensor/fiber orientation distribution fitting is described in detail in the supplementary text.

**Fig. 7.**
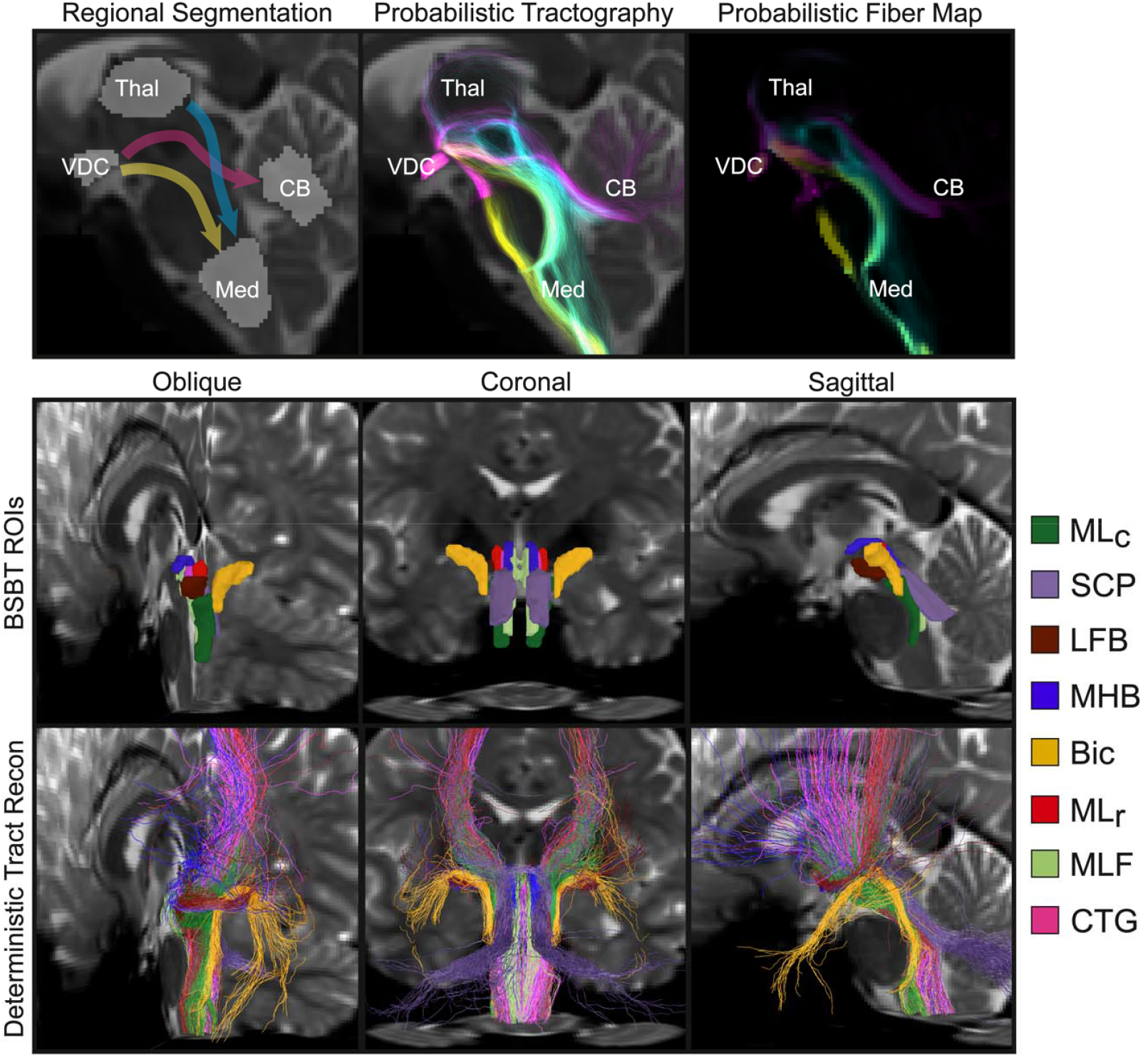
Probabilistic Fiber Map construction in the rostral brainstem. (top row) Tracts are seeded probabilistically with *MRtrix Tckgen* between four ROIs segmented with FreeSurfer that lie adjacent to the rostral brainstem: The Ventral Diencephalon (VDC), Thalamus (Thal), Cerebellar gray matter (CB) are segmented with *SynthSeg*, and the Medulla Oblongata (Med) was segmented with the FreeSurfer brainstem subfield segmentation package. The VDC, Thal, and CB masks in this figure are dilated by a 3-voxel kernel for better visualization. Individual streamline density maps are then created each ROI pair the tracts are seeded between. Deterministic tractography aids in visualizing tract reconstructions and reveals coherent bundles that are specific to each ROI pair, as well as multi-channel overlays from multiple streamline pairs. Probabilistic streamlines are then histogram-normalized and combined into a single 3-channel PFM, where each voxel contains a Cyan-Magenta-Yellow encoding. This encoding represents the corresponding voxels from the normalized streamline density maps for Thal-Med, VDC-CB, and VDC-Med ROI pairs respectively. **(bottom row)** Color-coded brainstem WM bundle ROIs segmented with BSBT in a representative *in vivo* dMRI scan are displayed over a low-b volume with corresponding deterministic streamlines. All streamlines intersected each brainstem WM bundle ROI along pre-specified ROI cross-sections. All deterministic tractography was performed with the *TrackVis* software package *(97)*.

### Convolutional Neural Network architecture

We used a U-Net CNN architecture previously levels, each containing 24 × 2 ^*n* −1^ is the level number), and two convolutional adapted for dMRI segmentation tasks for BSBT (89, 90). The CNN comprises five resolution features (where layers with 3 × 3 × 3 kernels and Exponential Linear Unit activation functions. The resolution is halved at every level using a max-pooling operator. The CNN takes in a five-channel input (low-b, FA, and PFM volumes). All input channels are resampled to 1mm isotropic resolution and cropped with dimensions of 64 × 64 × 64 voxels centered around the center-of-mass of the pontine ROI (obtained from the FreeSurfer brainstem subfield segmentation algorithm) during testing (and pre-cropped at training time). A schematic overview of the full CNN model is illustrated in Figure 8. Further details on CNN training and inference, including train-time data augmentation, can be found in the supplementary text.

**Fig. 8.**
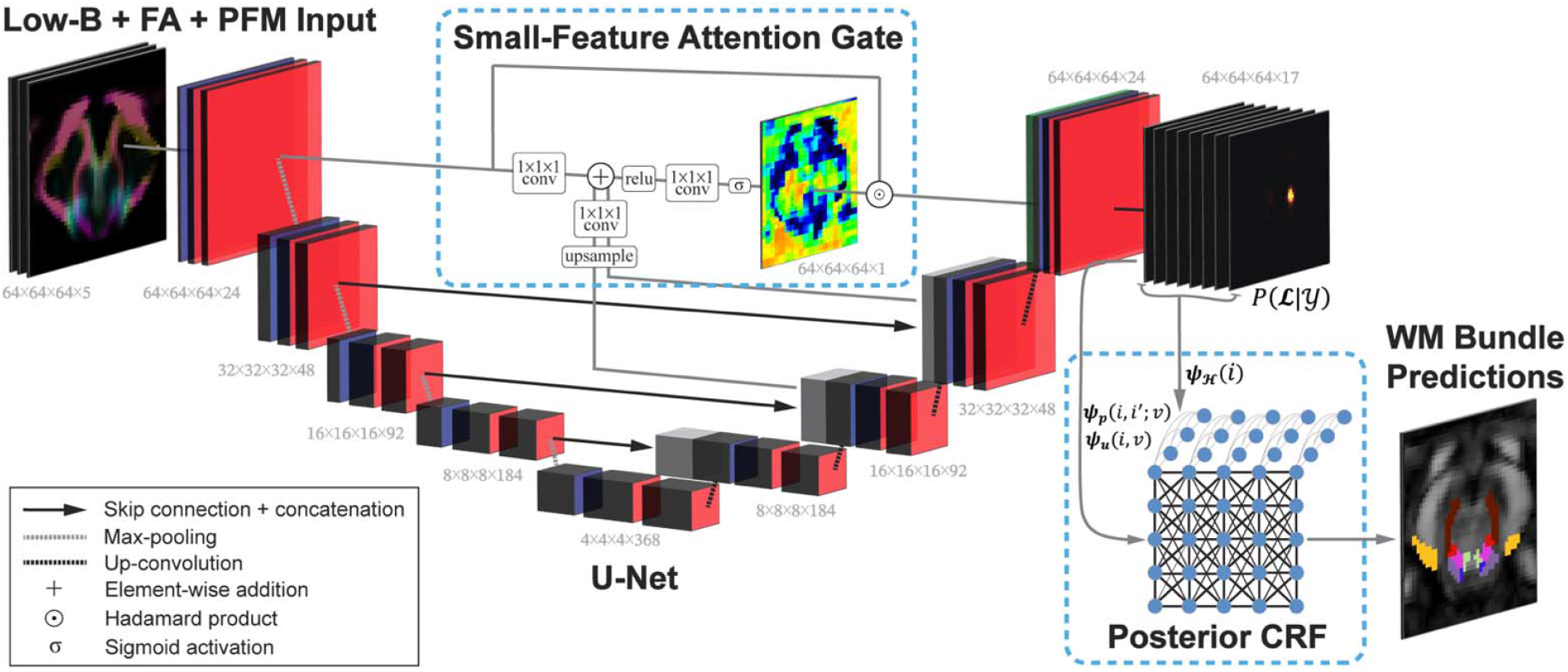
Overview of BSBT CNN architecture. A U-Net CNN modified with an attention gating mechanism placed on the three highest-resolution CNN layers takes 5 dMRI channels consisting of a low-b volume, FA volume, and corresponding PFM channel as an input. The *SoftMax* output of the CNN(*P*(***L***| ***Y))*** is then processed by the CRF with unary (ψ_*u*_), pairwise (*ψ* _*p*_), and label entropy (ψ_*p*_), potentials to output refined brainstem WM bundle segmentations.

Brainstem WM bundles constitute a small fraction of the total volume compared to surrounding brainstem structures. We enhance the CNN with an attention-gating mechanism, which are commonly used in biomedical image segmentation (91–94), to focus on and amplify small ROIs, thereby improving the model’s ability to capture subtle anatomical details constituting brainstem WM bundles. Normally, attention gates for a CNN encoder layer directly incorporate feature information from a single CNN decoder layer. We integrate inputs from multiple decoder layers to capture WM bundle feature information across spatial scales. Let the CNN encoder and decoder 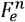 and 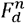 where *n* ∈ 1…*N* is the layer number of the CNN (ordered from fine/high-resolution to coarse/low-resolution) where 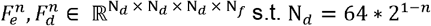 and N_*f*_ =24* 2^*n*−1^ We only implement gating for the skip connection between 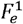 and 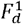. Second, we integrate gating signals from all decoder layers except for the coarsest layer (due to its overly-”blocky” feature representation through four voxels in each spatial dimension). Finally, rather than downsampling the input encoder layer 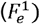 to the spatial resolution of gating signals from the decoder, we upsample all gating signals to the resolution 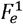 (and by extension 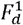) to preserve fine-scale features. These implementations aim to create a more interpretable gating model by additively combining multiple decoder receptive fields (i.e., gating signals) in one function. The gating signals are transformed with a hyperbolic tangent activation function prior being added to the incoming spatial layer for weight normalization, and to allow for both positive and negative outputs to mimic receptive field activation and suppression. We define the attention gating mechanism Φ*att* (·) as:

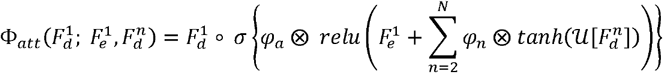

Where ∘ is the Hadamard product, ⊗ is the three-dimensional convolutional operator, φ is a 1 ×1 ×1 convolutional kernel, 𝒰 (·) is an upsampling operation for match the dimensionality 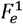 *relu*, (·) is a rectified linear unit, *tanh* {·} is the hyperbolic tangent function and σ {·} is a sigmoid activation function. The effects of each attention-gate modification are visualized with a representative *ex vivo* subject in Figure 9.

**Fig. 9.**
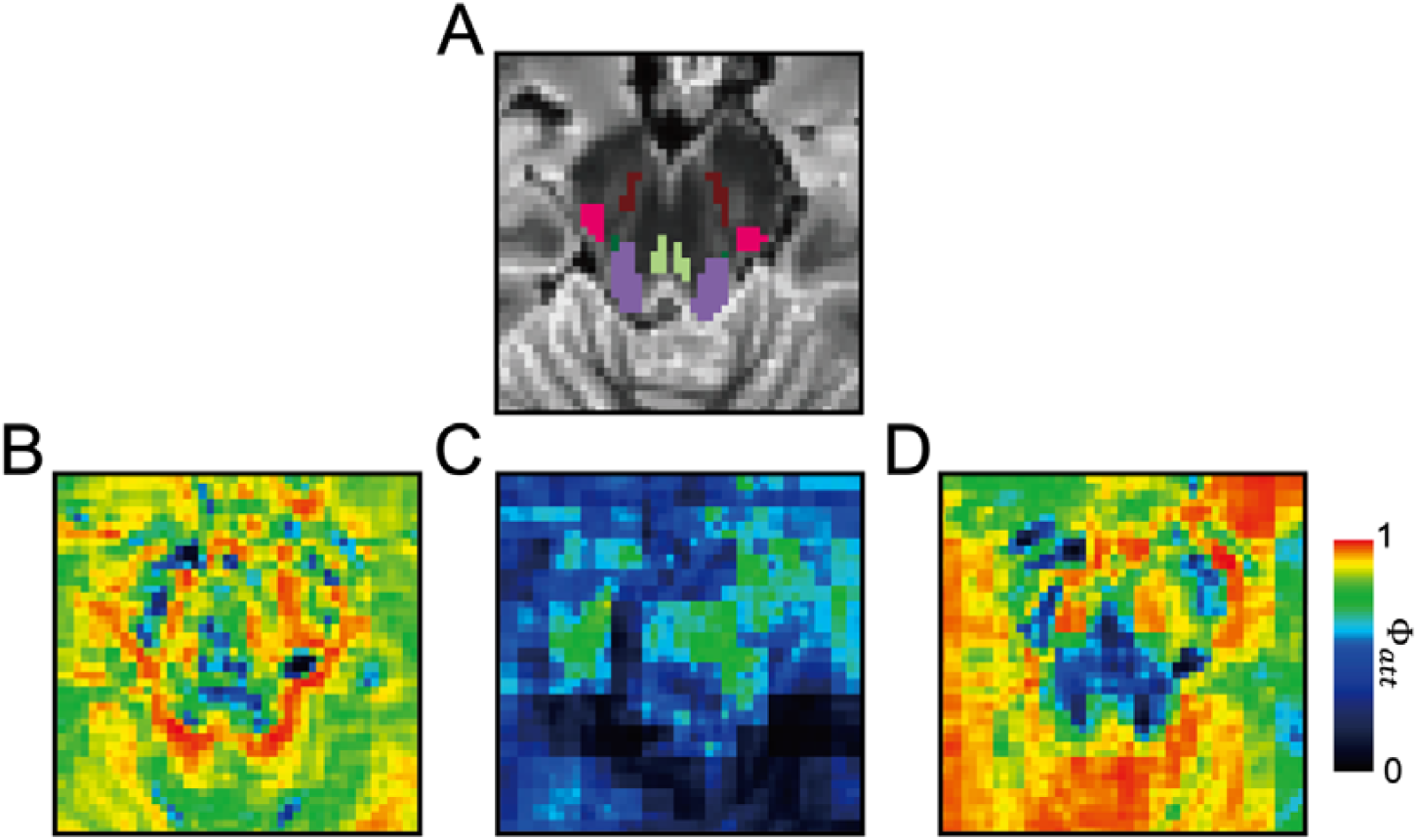
Attention-gating modifications for brainstem WM bundle segmentation. **(A)** Outputted attention map coefficients () are visualized from a representative *ex vivo* low-b image with overlayed BSBT segmentations. **(B)** Due to the small size and mixed sparsity of brainstem WM bundles, the base attention-gating module focuses well on individual WM bundle subregions but empirically tends to display poor coherence in regions with clustered WM bundles. **(C)** We additively combined features from each decoder layer (except the lowest-resolution layer) with the gating signal to incorporate information from multiple receptive fields and aid in detection of multi-label clusters. However, lower-resolution features tended to dominate and saturate attention coefficients, leading to block artifacts. **(D)** We subsequently incorporated hyperbolic tangent activation functions at the output of each feature and gating signal prior to addition, to aim for balanced mixing of receptive fields. Of note, both the standard and modified attention gating mechanisms displayed an ‘inverted’ gating signal in our CNN model, where attention coefficients were lower in WM bundle regions than in surrounding structures.

### Conditional random field design

The background label probabilities from the *SoftMax* CNN layer tends to dominate over and dilute the probabilities of small and thin brainstem WM bundle labels, leading to class imbalance (even with the use of a Dice score penalty during CNN training). This is more evident in domains differing from the training dataset, such as low-resolution *in vivo* and *ex vivo* dMRI. We employ a label probability map enhancement strategy for the *SoftMax* CNN layer with a semi-dense CRF (95). CRF refinement is employed to increase label probabilities near WM bundle label edges, which are often inpainted by the background label. Let the CNN input volume 𝒴 ∈ ℝ ^64 × 64 × 64 ×5^ consisting of *N* = 64^3^ voxels in a uniform grid, a corresponding ground-truth label map ℒ ∈ ℝ^*N*×*V*^ consisting of *V* labes, such that, for a spatial location (i.e., voxel) 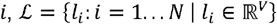 We infer the refined label probabilities *P*(ℒ | 𝒴) with the CNN, which are originally approximated with the *SoftMax* output *S* ∈ ℝ^*N*×*V*^. The joint posterior probability distribution can be written as:

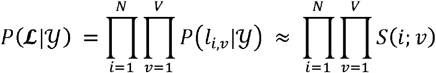

For CRF refinement, we reformulate *P*(ℒ | 𝒴) as a Gibbs distribution with the CRF energy functional E(ℒ;.) such that:

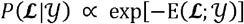

We model E(ℒ;.) as a linear combination of the negative log-likelihood ψ_*u*_ log-pairwise potential ψ_*p*_ and label entropy regularizer ψ _*ℋ*_ (95, 96). We refine *P*(ℒ | 𝒴) to better capture label interactions through Maximum a Posteriori inference, which is equivalent to minimizing E(ℒ;.) to determine the updated labels ℒ_CRF:_

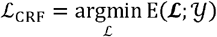

ψ_*u*_ (the negative log-likelihood of the label probabilities) and ψ _*ℋ*_ are defined voxel-wise and separately for each label (where we assign *v* = 1as the background label) as:

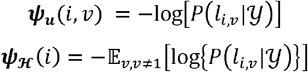

ψ_*ℋ*_ regularizes certainty (i.e., distinctness) for foreground labels without background label interference. This preserves WM bundle label structure by maintaining sharp boundaries between adjacent labels, while allowing label probabilities to dominate in regions of background inpainting (i.e., in regions of anatomically-distinct foreground with uniformly dominant background posteriors). We decompose **ψ**_***p***_ into spatial 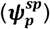 and intensity 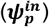 components, in order to sensitize the CRF to intensity fluctuations in adjacent voxels. We define **ψ**_***p***_ or the voxel pair (i,i′) around a local neighbourhood 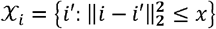 as:

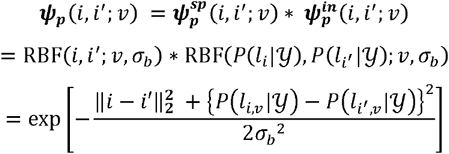

where‖ · ‖ _2_ is the Euclidean norm, and RBF(·; σ) is a Radial Basis Function operator parameterized by σ We assume that 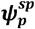 and 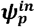 possess equal importance such that RBF(·; σ) is parameterized by a common σ_*b*_ To exploit the additive nature of potentials in log-space, we express the total CRF energy penalty used for computation as:

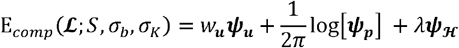

where we collapse the pairwise indicator function and approximate the 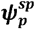 RBF operator by convolving *S* with an RBF kernel 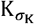 parameterized by σ_K_ which encapsulates the neighborhood size of the CRF. This convolution aids in GPU acceleration, decreases run-time, and reduces the log-pairwise potential to:

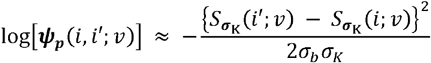

where:

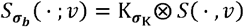

The explicit form of the CRF energy penalty used for computation(E_*comp*_) and applied to *S* can therefore be expressed as:

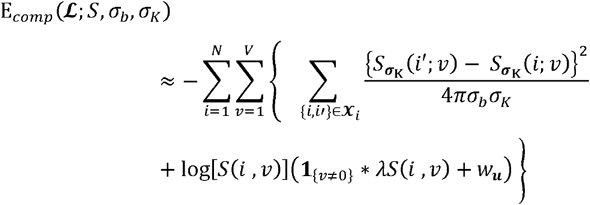

Where **1** {_·_} is the indicator function. We set the σ_K_ to five voxels, and neighborhood span of **χ**_*j*_ to three voxels to permit faster computation times. The CRF energy penalty above is refined, but not solved until convergence to a minimum, with an iterative mean-field approximation method over a small number of iterations with a fixed run-time to avoid overfitting.

## Data Availability

All code used for probabilistic mapping, segmentation and statistical analysis can be found at https://github.com/markolchanyi/BSBT. All digitized histopathological sections can be found on the Biolucida viewer (https://histopath.nmr.mgh.harvard.edu/images/?page=images&selectionType=collection&selectionId=50). All diffusion sequences, FLASH sequences and representative PFM reconstructions for the ex vivo brain specimens used in our analysis can be found on OpenNeuro (https://openneuro.org/datasets/ds006001). Minimally preprocessed diffusion and structural sequences for ADNI and PPMI subjects can be found on the University of Southern California Image and Data Archive (IDA) (https://ida.loni.usc.edu). Minimally preprocessed diffusion and structural sequences for HCP subjects can be found on ConnectomeDB (https://db.humanconnectome.org/).

https://histopath.nmr.mgh.harvard.edu/images/?page=images&selectionType=collection&selectionId=50

https://openneuro.org/datasets/ds006001

## Acknowledgments

Data collection and sharing for this AD analysis in this project was funded by the Alzheimer’s Disease Neuroimaging Initiative (ADNI) (National Institutes of Health Grant U01 AG024904) and DOD ADNI (Department of Defense award number W81XWH-12-2-0012). ADNI is funded by the National Institute on Aging, the National Institute of Biomedical Imaging and Bioengineering, and through generous contributions from the following: AbbVie, Alzheimer’s Association; Alzheimer’s Drug Discovery Foundation; Araclon Biotech; BioClinica, Inc.; Biogen; Bristol-Myers Squibb Company; CereSpir, Inc.; Cogstate; Eisai Inc.; Elan Pharmaceuticals, Inc.; Eli Lilly and Company; EuroImmun; F. Hoffmann-La Roche Ltd and its affiliated company Genentech, Inc.; Fujirebio; GE Healthcare; IXICO Ltd.; Janssen Alzheimer Immunotherapy Research & Development, LLC.; Johnson & Johnson Pharmaceutical Research & Development LLC.; Lumosity; Lundbeck; Merck & Co., Inc.; Meso Scale Diagnostics, LLC.; NeuroRx Research; Neurotrack Technologies; Novartis Pharmaceuticals Corporation; Pfizer Inc.; Piramal Imaging; Servier; Takeda Pharmaceutical Company; and Transition Therapeutics. The Canadian Institutes of Health Research is providing funds to support ADNI clinical sites in Canada. Private sector contributions are facilitated by the Foundation for the National Institutes of Health (www.fnih.org). The grantee organization is the Northern California Institute for Research and Education, and the study is coordinated by the Alzheimer’s Therapeutic Research Institute at the University of Southern California. ADNI data are disseminated by the Laboratory for Neuro Imaging at the University of Southern California. Data were also provided [in part] by the Human Connectome Project, WU-Minn Consortium (Principal Investigators: David Van Essen and Kamil Ugurbil; 1U54MH091657) funded by the 16 NIH Institutes and Centers that support the NIH Blueprint for Neuroscience Research; and by the McDonnell Center for Systems Neuroscience at Washington University. Data used [in part] in the preparation of this article was obtained on 2024-12-16 from the Parkinson’s Progression Markers Initiative (PPMI) database (www.ppmi-info.org/access-data-specimens/download-data), RRID:SCR_006431. For up-to-date information on the study, visit www.ppmi-info.org. PPMI – a public-private partnership – is funded by the Michael J. Fox Foundation for Parkinson’s Research and funding partners, including 4D Pharma, Abbvie, AcureX, Allergan, Amathus Therapeutics, Aligning Science Across Parkinson’s, AskBio, Avid Radiopharmaceuticals, BIAL, BioArctic, Biogen, Biohaven, BioLegend, BlueRock Therapeutics, Bristol-Myers Squibb, Calico Labs, Capsida Biotherapeutics, Celgene, Cerevel Therapeutics, Coave Therapeutics, DaCapo Brainscience, Denali, Edmond J. Safra Foundation, Eli Lilly, Gain Therapeutics, GE HealthCare, Genentech, GSK, Golub Capital, Handl Therapeutics, Insitro, Jazz Pharmaceuticals, Johnson & Johnson Innovative Medicine, Lundbeck, Merck, Meso Scale Discovery, Mission Therapeutics, Neurocrine Biosciences, Neuron23, Neuropore, Pfizer, Piramal, Prevail Therapeutics, Roche, Sanofi, Servier, Sun Pharma Advanced Research Company, Takeda, Teva, UCB, Vanqua Bio, Verily, Voyager Therapeutics, The Weston Family Foundation and Yumanity Therapeutics.

We are immensely thankful to each family for the donation of the brains of their loved ones, as well as to the individuals who donated their brains for this study and the overall advancement of medical science. We thank Michelle Siciliano for help in processing of human brain specimens and Kathryn Regan at Veranex, Inc. for assistance with blocking, sectioning and immunostaining of histological data.

## Notes

### Competing Interest Statement

The authors have declared no competing interest.

### Funding Statement

This work was supported by the National Institutes of Health Directors Office (DP2hd101400), National Institute for Neurological Disorders and Stroke (R21NS109627, K23NS094538, 1R21NS138995, U01NS086625, R01NS0525851, R21NS072652, R01NS070963, R01NS083534, uh3NS095554, U24NS10059103, R01NS105820, and RF1NS115268), National Institute for Biomedical Imaging and Bioengineering (P41EB030006, R01EB023281, R01EB006758, R21EB018907, R01EB019956, and 1R01EB031114), National Institute of Child Health and Human Development (R01hd102616 and R21hd095338), National Institute on Deafness and Other Communication Disorders (R21dc015888), National Institute on Aging (1R01AG070988, R21AG082082, R01AG064027, R01AG008122, 1RF1AG080371, R01AG016495, and R01AG070988), National Institute on Mental Health (1RF1MH123195, UM1MH130981, R01MH123195, 1UM1MH130981, R01MH121885, and RF1MH123195), Brain Initiative Cell Census Network (U01MH117023 and UM1MH130981), James S. McDonnell Foundation, American Academy of Neurology/American Brain Foundation, Center for Integration of Medicine and Innovative Technology (Boston, MA), Rappaport Foundation, American SidS Institute, Chen Institute MGH Research Scholar Award, and the MIT/MGH Brain Arousal State Control Innovation Center (BAScic) project. Additional support was provided by the Blueprint for Neuroscience Research (U01MH093765), part of the multi-institutional Human Connectome Project. This research was also made possible by the resources provided by NIH grant P41rr014075 and shared instrumentation grants S10rr023401, S10rr019307, and S10rr023043. Computational resources for this research were generously provided by the Massachusetts Life Sciences Center (www.masslifesciences.com/). MDO received funding from the MIT-Takeda fellowship and the MIT IMES fellowship. DRS received funding from the Swiss National Science Foundation (SNSF) [P500PM_210834]. ASA received funding from the American Heart Association-Tedys Team postdoctoral fellowship award (24POST1023166).

### Author Declarations

IRB of Massachusetts General Hospital gave ethical approval for this work

