## Supporting Material for "Probabilistic Mapping and Automated Segmentation of Human Brainstem White Matter Bundles"

**This PDF file includes:**

Supporting text

Figures S1 to S15

Tables S1 to S3

Legend for Dataset S1

SI References

Supporting Information Text

**Annotation protocol for brainstem white matter bundles**

**Pontine (caudal) division of the Medial Lemniscus (MLc).** We aimed to have the neuroanatomic borders and positioning of our MLc annotations be consistent with the pontine and caudal-midbrain subdivisions of the "ml" (medical lemniscus) ROI in the Paxinos Atlas of Human Brainstem (1). More specifically, the caudal extent of the BSBT MLc annotation terminated at plate #8.32 (obex +15mm), and the rostral extent terminated at plate #8.56 (obex +39mm).

**Mesencephalic (rostral) division of the Medial Lemniscus (MLr).** We aimed to have the neuroanatomic borders and positioning of our MLc annotations be consistent with the pontine and rostral-midbrain subdivision of the "ml" (medical lemniscus) ROI in the Paxinos Atlas of Human Brainstem. The BSBT MLr annotation follows in rough-continuity with our MLc annotation, the caudal extent of which begins at plate #8.57 (obex +40mm) where the Paxinos ml ROI becomes adjacent to the dorso-lateral border of the red nucleus. The MLr annotation then follows the ml ROI to the rostral-most Paxinos axial plate (#8.64, obex +47mm), then roughly 2mm rostrally to the axial plane corresponding with the top of the red nuclei. Of note, we do not segment the lateral "wings" of the MLr (as can be seen in the Paxinos atlas), do to their comparatively small cross-sectional area. The MLr is also observed to be a distinctly hypo-intense region extending from the dorso-lateral margin of the red nucleus in the ultra-high resolution FLASH sequence.

**Superior Cerebellar Peduncle (SCP).** We aimed to have the neuroanatomic borders and positioning of our SCP annotations be consistent with the pontine subdivisions of the "scp" (superior cerebellar peduncle) ROI in the Paxinos Atlas of Human Brainstem. The rostral extent of the BSBT SCP annotation terminated at the plane of initiation of the SCP decussations (xSCP) located at plate #8.48 (obex +31mm). The caudal extent of our SCP annotation extends roughly 5mm caudal to, and 6mm dorsal to the most caudal Paxinos plate with an SCP ROI (plate #8.35, obex +18mm) into the hemispheric white matter of the cerebelli.

**Brachium of the Inferior Colliculus (Bic).** We aimed to have the neuroanatomic borders and positioning of our Bic annotations be consistent with the pontine subdivisions of the "bic" (brachium of the inferior colliculus) ROI in the Paxinos Atlas of Human Brainstem. The caudal extent of the BSBT Bic annotation coincided with the most caudal plate with a bic ROI in the Paxinos atlas, located at the ponto-mesencephalic junction (plate #8.52, obex +35mm). Our BIC annotation follows the neuroanatomic borders of the Paxinos bic ROI to its rostral termination plane at plate #8.60 (obex +43mm). Our annotation then extends roughly 5mm rostrally to this plate, first along the dorso-lateral borders of the medial and ventral subunits of the mediate geniculate nucleus (MGM/MGV) and the posterior intralaminar thalamic nucleus (PIL), then along the dorsal borders of the medial geniculate nucleus (MG) and posterior ventromedial thalamic nucleus (PVM).

**Medial Longitudinal Fasciculus (MLF).** We aimed to have the neuroanatomic borders and positioning of our Bic annotations be consistent with the pontine subdivisions of the "mlf" (medial longitudinal fasciculus) ROI in the Paxinos Atlas of Human Brainstem. The caudal-most extent of the BSBT MLF annotation is located at plate #8.34 (obex +mm), where the MLF starts splaying laterally from the pontine midline. Our MLF annotation then follows the Paxinos mlf ROI to the rostral-most Paxinos axial plate (#8.64, obex +47mm). The MLF is also observed to be a distinct hypo-intense region ventral to the periaqueductal gray (Paxinos: PAG) in the midbrain and lateral to the median raphé nucleus (Paxinos: MnR) the ultra-high resolution FLASH sequence.

**Central Tegmental Tract (CTG).** We aimed to have the neuroanatomic borders and positioning of our CTG annotations be consistent with the midbrain subdivision of the "ctg" (central tegmental tract) ROI in the Paxinos Atlas of Human Brainstem. The caudal-most extent the BSBT begins roughly 2mm rostral to the ponto-mesencephalic junction (plate 8.54), and follows the Paxinos ctg ROI along the mid-dorsal border of the red nuclei to the rostral-most Paxinos axial plate (#8.64, obex +47mm). The CTG is also observed to be a weakly hypo-intense sagittal band medial and dorsal to the margins of the red nucleus in the ultra-high resolution FLASH sequence.

**Lateral Forebrain Bundle (LFB) and Mesencephalic Homeostatic bundle (MHB)**. Neither the LFB nor the MHB ROIs are included as part of the Paxinos Atlas of the Human Brainstem. However, the morphology and connectivity of both bundles has been outlined in prior literature (1*4*). The LFB was observed as a distinct WM bundle with high streamline intensities in the third PFM channel and pronounced myelination in corresponding HE-LFB sections. The LFB was delineated as located proximal to the ventrolateral margin or the Red Nuclei. The MHB revealed Ventral Diencephalon-based streamline intensities in the PFM channel and high white matter density in corresponding histological sections that were along the superior/medial to the red nuclei and along the inferior\lateral margin of the third ventricle in the rostral extent, and ventrolateral to the Periaqueductal Grey in the caudal extent. Post-hoc deterministic streamline tractography with TrackVis software version 0.6.2 (2) (of streamlines seeded from ROIs annotated over the LFB candidate region produced coherent bundles that closely resembled prior LFB reconstructions (3) (see Figure S2). Deterministic streamline tractography of streamlines seeded at the MHB ROI produced coherent bundles that resembled MHB reconstructions at the caudal extent from prior literature (3) and merged with the Medial Forebrain Bundle at the rostral extent (see Figure S3). The MHB is also observed as a weakly hypo-intense sagittal band extending rostrally and medially to the red nuclei in the ultra-high resolution FLASH sequence.

**MRI datasets**

This supplementary section provides dataset information, scanning parameters, inclusion/exclusion criteria, and notes on quality issues for all utilized dMRI, structural MRI, and histology data. Further details on individual subject parameters, including subject IDs (if available), age, sex, and inclusion/exclusion status for analysis can be found in the supplementary Excel file. Exclusion criteria for all segmentations in data used for test-retest and clinical analysis (i.e., HCP test-retest, AD, PD, and TBI data) were (1) one or more missing segmentation labels or (2) sparse/grossly-incomplete segmentations less than ~20 voxels in volume. These exclusion criteria were applied to BSBT, *TractSeg* WM bundles, and *SynthSeg* hemispheric gray matter/brainstem masks. All excluded segmentations were visually inspected and confirmed. No exclusion criteria were applied for Dice score, as to fairly interpret accuracy/ablation analysis. WM bundles with missing segmentation labels within a subject were excluded from HD calculation, as a lack of a label leads to non-computable HD values.

**Human Connectome Project data.** As part of our analysis, we used healthy subject dMRI and T1 sequence data from the HCP dataset (4, 5). We chose random subsets of 30 subjects for training of the CNN models, 8 subjects for offline validation of CNN training, and 10 subjects for accuracy/ablation analysis. Furthermore, we used 46 subjects who underwent two separate scanning sessions, which were included as part of the "WU-Minn HCP Retest Data" group, for our test-retest analysis. Two of the 46 subjects were excluded from analysis due to not undergoing dMRI scanning in either the first or follow-up scanning sessions. Four additional subjects were excluded from test-retest analysis as they were used for CNN training prior to any validation studies. dMRI volumes were acquired on a Siemens CommectomS scanner (Siemens Medical Solutions, Erlangen, Germany) at 3 Tesla (TR: 5520 ms, TE: 89.50 ms, flip angle: 78°) and contain 110 diffusion encoding directions split into three shells (b=1000, 2000, and 3000 s/mm^2^) and 18 low-b (b=0 s/mm^2^) volumes at 1.25 mm isotropic resolution. Further information can be found on the [Human Connectome Project website](https://humanconnectome.org/study/hcp-young-adult/document/1200-subjects-data-release). T1 volumes used to aid in manual annotation were acquired using a Magnetization Prepared - RApid Gradient Echo (MP-RAGE) sequence at 0.7 mm isotropic resolution (TR: 2400 ms, TE: 2.14 ms, flip angle: 8°).

**Ex Vivo brain specimens.** We used seven *ex vivo* brain specimens for analysis. Each specimen was donated from individuals who died of non-neurological causes, had no history of neurologic disease, and displayed no gross abnormalities upon post-mortem brain examination by a neuropathologist. For each brain specimen, we obtained written informed consent from family members designated as surrogate decision makers, as per accordance with a protocol approved by the Mass General Brigham Institutional Review Board. Further details on each brain specimen can be found in Table S2. All seven specimens were used for accuracy/ablation analysis. Two of the seven specimens were used along with guidance from corresponding histological sections (see "Histology data" in the supplementary text) for determining locations and morphology of brainstem WM bundles suitable for segmentation. Each specimen was scanned on a large-bore 3 Tesla Siemens Tim Trio scanner (Siemens Medical Solutions, Erlangen, Germany) in a custom 32-channel head coil. dMRI was acquired with a Steady State Free-Precession sequence (TR: 38 ms, TE: 23 ms, flip angle: 60°) with a single shell (effective b=3773 s/mm^2^) with 90 diffusion encoding directions and 12 low-b volumes at 750 μm isotropic spatial resolution. Corresponding ultra-high resolution structural volumes for each specimen were acquired at 200μm isotropic spatial resolution in a whole-body 7 Tesla Siemens Magnetom scanner (Siemens Medical Solutions, Erlangen, Germany) using the aforementioned custom head coil and a Fast Low-Angle SHot (FLASH) sequence (TR: 40 ms, TE: 14.2 ms, flip angle: 20°) acquired at 550 μm isotropic spatial resolution (6). For specimen S3 in table S1, the FLASH sequence was acquired at 550 μm isotropic spatial resolution

**Alzheimer's Disease Neuroimaging Initiative data.** All dMRI and structural data used in the analysis described below was obtained from the Alzheimer's Disease Neuroimaging Initiative (ADNI) database (7, 8) (adni.loni.usc.edu). The ADNI was launched in 2003 as a public-private partnership, led by Principal Investigator Michael W. Weiner, MD. The original goal of ADNI was to test whether serial magnetic resonance imaging, positron emission tomography, other biological markers, and clinical and neuropsychological assessment can be combined to measure the progression of mild cognitive impairment and early AD. We analyzed a cognitively normal (control) group consisting of 138 subjects, and a clinical group consisting of 113 subjects who were diagnosed with either AD or early/late-stage MCI (AD/MCI group). We grouped together AD and MCI groups for our analysis due to the comparatively-low number of AD subjects with suitable dMRI acquisitions (n = 25). All subject data was acquired from the ADNI3 dataset. Seven subjects from the control group and 16 subjects from the AD/MCI group were excluded from analysis. The AD/MCI and control groups (post-exclusion) were age- and sex-matched, with Wilcoxon rank sum p-value of $0.31$ for age and a Chi-Squared p-value of $0.25$ for sex. We used all subjects from both groups for clinical analysis and randomly chose a subset of 10 subjects from the control group to manually annotate for accuracy/ablation analysis. All subjects used for our analysis were scanned with an axial "ADNI3-Basic" sequence (TR: 7200 ms, TE: 56 ms, flip angle: 90°) with a single shell (b=1000 s/mm^2^) with 48 diffusion-encoding directions and 7 low-b volumes at 2mm isotropic spatial resolution. In the 10 subjects used for accuracy/ablation analysis, we also utilized accelerated MP-RAGE volumes (TR: 2300 ms, TE: 3 ms, variable flip angle) scanned at a 1-1.1mm in-plane resolution and 1-1.2mm slice thickness to aid in manual annotation of brainstem WM bundles. Further information on subject and acquision information can be found at [adni.loni.usc.edu](https://adni.loni.usc.edu/).

**Parkinson's Progression Biomarkers Initiative data.** We analyzed a cognitively-normal (control) group consisting of 60 subjects and a clinical (PD) group consisting of 134 subjects diagnosed with PD. All subject data was acquired from the publicly-available PPMI dataset (9). Each subject from the PD group underwent two scanning sessions, one baseline scan at time of diagnosis and a 2YFU scan. Eight subjects from the control group and 62 subjects from the PD group were excluded from analysis. A further exclusion criterion specific to PD group was successful segmentation of all WM bundles and gray matter regions in both the baseline and 2YFU scanning sessions. The control and PD group at the baseline groups (post-exclusion) were age- and sex-matched, with Wilcoxon rank sum p-value of $0.85$ for age and a Chi-Squared p-value of $0.67$ for sex. For consistency, we chose dMRI data from all subjects that was acquired with a gated 2D Echo-Planar Imaging dMRI sequence on a 3 Tesla Siemens TimTrio scanner (Siemens Medical Solutions, Erlangen, Germany) (TR: 500-1000 ms, TE: 88 ms, flip angle: 90°) with a single shell (b=1000 s/mm^2^) containing 54 diffusion-encoding directions and 1 acquired low-b volume. Due to the comparably low signal-to-noise ratios of the dMRI scans, as shown in Figure S15, we pre-smoothed all FA maps with a Gaussian kernel with a standard deviation of 1mm. Of note, the comparatively low signal-to-noise ratio, and potentially the sparce segmentations/high segmentation failure rate, was likely due to a single low-b volume contained in the dMRI sequence, in turn limiting the effectiveness of dMRI preprocessing and denoising. Further information on subject and acquision information can be found at [www.ppmi-info.org](https://www.ppmi-info.org/).

**Traumatic Brain Imaging dataset.** We analyzed 33 control subjects with no prior history of neurologic or cardiovascular disease, and 18 acute TBI patients enrolled in an ongoing TBI study (10) (ClinicalTrials.gov: NCT03504709). Sixteen of the patients (p1-16, see Table S2) were enrolled in the pilot phase of the study. Informed consent was obtained directly from control subjects and via surrogate decision makers for the acute TBI patients in accordance with protocol approved by the Mass General Brigham Institutional Review Board. Four control subjects and one TBI patient were excluded from analysis. The control and TBI groups (post-exclusion) were age- and sex-matched, with Wilcoxon rank sum p-value of $0.39$ for age and a Chi-Squared p-value of $0.35$ for sex. All subjects were scanned in a 3 Tesla Siemens Skyra scanner (Siemens Medical Solutions, Erlangen, Germany) in a custom 32-channel head coil an Echo-Planar Imaging dMRI sequence (TR: 13700 ms , TE: 98 ms) with a single shell (b=2000 s/mm^2^) and 10 low-b volumes at 2mm isotropic spatial resolution. Nine patients and 4 control subjects were scanned with the same protocol and parameters, but with the addition of Simultaneous Multi-Slice acceleration (11). MP-RAGE sequences (TR: 2530 ms, TE: 1.69 ms, flip angle: 7°) were acquired for all subjects on the same scanner at a 1mm isotropic spatial resolution. Finally, 3D SWIs (TR: 30 ms, TE: 20 ms, flip angle: 15°) were acquired for each subject at 0.86 × 0.86 × 1.8 mm spatial resolution. SWI scans from subjects P5 and P16 (overviewed in Table S2) were excluded from overlap analysis due to the presence of significant motion artifacts (12).

**Histology data.** The brainstems of two of the seven *ex vivo* specimens were cut into three blocks and the rostral blocks (midbrain and pons) were serially sectioned with slice thicknesses of 10 μm with a microtome (LEICA RM2255 microtome, Leica Microsystems, Buffalo Grove, IL, USA). Every 50th section (i.e., every 500 μm) was stained with Hematoxylin/Eosin and counterstained with Luxol Fast Blue (HE-LFB) in accordance with a previously defined histopathology/immunohistochemistry protocol (13). All histological sections were then digitized with the NanoZoomer S60 Digital Slide Scanner (Hamamatsu Photonics). Digitized whole-slides were postprocessed with custom scripts for white balance correction and contrast enhancement. All slides were then converted to a JPEG format and published on the Biolucida platform at [histopath.nmr.mgh.harvard.edu](https://histopath.nmr.mgh.harvard.edu/images/?page=images&selectionType=collection&selectionId=45).

**Diffusion MRI preprocessing for Probabilistic Fiber Map construction**

Each dMRI scan underwent standard preprocessing which included Marchenko-Pastur PCA denoising (using the MRtrix *dwidenoise* command (14)), followed by brain mask extraction, motion correction, susceptibility-induced distortion correction, and eddy current correction using FSL commands all wrapped by the *dwifslpreproc* command (15). ANTs N4-based bias field correction was then performed using the MRtrix *dwibiascorrect* command. The preprocessed dMRI volume is resampled to a 1mm isotropic resolution with bicubic interpolation in order to avoid fitting tractography parameters dependent on voxel size. FA was derived from a tensor fitting with MRtrix *dwi2tensor* and *tensor2metric* commands. We obtain the WM response function, which estimates the expected signal for an oriented WM bundle, with either the *tournier* algorithm for the single-shell volumes or the *dhollander* algorithm for the multi-shell volumes (16, 17). We then calculate fiber orientation distribution functions for each voxel with constrained spherical deconvolution with the WM response function serving a symmetric starting kernels using the MRtrix *dwi2fod* command.

**Hemorrhagic lesion localization and registration in Susceptibility-Weighted Images**

To rigorously benchmark BSBT, we enhanced the discriminatory power of the *TractSeg* LDA classifier by selecting WM bundles with the greatest spatial overlap with hemorrhagic lesions for ROC analysis. We also evaluated hemorrhagic lesion overlap with brainstem WM bundles to assess linkages between FA reduction and hemorrhagic lesioning. For lesion localization, we manually traced hypointense hemorrhagic regions in SWI, where changes in magnetic susceptibility creates hypointense SWI contrast in regions of hemorrhage that contain paramagnetic blood derivatives such as deoxyhemoglobin and hemosiderin (18, 19). Lesions were traced in SWI scans from 15 of the 17 TBI subjects; two of the 17 SWIs were not used due to significant motion artifacts (12). We translated lesion ROIs from SWI into dMRI space with affine image registration using ANTs (20) to directly compare with WM ROIs. Degrees of spatial overlap with both supra-tentorial and brainstem WM bundles were calculated as a Dice score. We chose 15 *TractSeg* WM bundles with the highest lesion overlap Dice scores as LDA classifier features. M.D.O manually traced all lesions in SWI. An expert neuropathologist, H.C.K, corrected and confirmed all SWI annotations. Each SWI volume was then registered to the corresponding low-b dMRI volume with an ANTs affine transformation model. Traced lesion masks were then propagated with the affine matrix from SWI space to dMRI space. The lesion localization and registration process are illustrated in Figure S11.

**Manual brainstem white matter bundle annotation for neural network training and accuracy/ablation analysis.**

We chose brainstem WM bundles to segment by induction. We first identified PFM contrast in (lower-resolution) *in vivo* dMRI volumes from HCP subjects that morphologically resembled known WM bundles. We then located analogous PFM regions in (higher-resolution) *ex vivo* dMRI volumes. To visually validate brainstem WM bundles in *ex vivo* space, we cross-referenced the anatomic locations and morphologies of the aforementioned PFM regions in dMRI space to HE-LFB sections provided for two of the seven *ex vivo* brain specimens which underwent histological sectioning. Specifically, this cross-referencing aided in determination of correspondence between true myelinated WM bundles and PFM contrast (Figure 1). Each WM bundle was identified with the aid of the Paxinos atlas of the human brainstem (1). We proceeded to label all chosen WM bundles in the PFM volume for accuracy/ablation analysis in these two brain specimens. We referred to the FLASH volume as an intermediate between histology and dMRI, which helped to determined axial locations of WM bundle margins in HE-LFB sections, as well as neuroanatomic contrast boundaries common to both histological and MRI space. With this top-down approach to WM bundle identification and localization, we facilitate the generalizability of BSBT in both *ex vivo* and *in vivo* as well as low- and high resolution dMRI domains. We labelled brainstem WM bundles on the PFM volume in the five *ex vivo* control brain specimens that did not undergo histological sectioning in the PFM volume with guidance from the corresponding FLASH volumes, as they possessed enough neuroanatomic contrast for accurate delineation of all bundles (see supplementary text and Figure 1D). For *in vivo* dMRI sequences (HCP and ADNI3 control subjects) used for accuracy/ablation analysis, we labelled brainstem WM bundles on the PFM volume with guidance from corresponding T1 scans. All annotation was performed with indirect guidance from the Paxinos atlas. M.D.O performed all manual labelling. An expert neuropathologist (H.C.K) corrected and confirmed label validity.

**Data augmentation and neural network training**

We trained the CNN for 480 epochs (1000 forwards and backwards passes of the network per epoch) on 30 HCP subjects with manually-labelled brainstem WM bundles. We used 8 manually-labelled HCP subjects for offline validation, which we evaluated at 15-epoch intervals (i.e., when intermediate models are saved). We trained the CNN with an ADAM optimizer (21) in two stages: a cross-entropy loss for a burn-in period of five epochs, followed by Dice loss until convergence. We used a cross-entropy burn-in to stabilize CNN model parameters and facilitate faster convergence during early training iterations. We then used Dice loss to mitigate class imbalance from variable sized brainstem WM bundles. We did not apply learning rate scheduling to training. We used aggressive augmentation to generalize the CNN model across dMRI modalities, resolutions, and signal-to-noise ratio levels. We pre-computed PFM volumes prior to augmentation in order to avoid re-running tractography at train-time. Apart from shell dropout randomization, we applied all augmentation strategies concurrently to the low-b, FA, and PFM channels. Our general augmentation strategies were 1: *Shell dropout*: In addition to the original multi-shell HCP dMRI data, we included separate FA and PFM reconstructions for each of the three provided shells (b=1000, 2000, and 3000 s/mm^2^) in the training dataset, as to mimic lower angular resolution and lower-contrast dMRI. 2: *Rotation and flipping*: We sampled rotation angles along the right, anterior and superior axes from a uniform distribution clipped between -25 and 25°. We performed left-right flipping to avoid lateralization effects. 3: *Intensity randomization and noise injection*: We randomly scaled intensities by $\pm$ 30%, performed histogram randomization, and added gaussian noise with a 0.01 standard deviation to each input channels independently, as to mimic WM tissue changes in neurological disease, high physiological noise (22), inhomogeneous fixation and reduced diffusivity in *ex vivo* dMRI (23). 4: *Resampling and deformation*: To address variability in resolution across dMRI domains and mimic partial voluming, we resample each channel with a gaussian kernel to a resolution of 2mm along each axis. 5: *Deformation.* To mimic local WM bundle morphologies affected by structural pathology, such as mass-effect from nearby lesions, we apply aggressive deformation and spatial resampling. Each CNN input channel is deformed using a basis-spline model with randomized gridding, then trilinearly interpolated.

**Neural network inference**

We calculated the posterior layer of the CNN before CRF refinement by averaging the CNN *SoftMax* output for an original and left/right-flipped input. Total inference time for a single HCP subject was recorded to be 2 seconds for a single CNN pass and 34 seconds for CRF processing over a 100-run average. Inference was performed with 8 AMD Ryzen Threadripper PRO 5995WX CPUs and a single NVIDIA RTX A6000 GPU.

**Region of Interest/segmentation comparison metrics**

We use two similarity metrics for assessment of BSBT segmentation accuracy with respect to ground-truth manual annotations: Dice score and average HD. For a segmentation mask $SM$ and ground truth mask $GT$, the Dice score and HD are defined as:

Dice score = $\frac{2 \times\left| SM \cap GT \right|}{\left| SM \right| + \left| GT \right|}$

HD $=\frac{\max_{s\in SM} \min_{g\in GT} \left\| s-g \right\|_{2} +\max_{g\in GT} \min_{s\in SM} \left\| g-s \right\|_{2}}{2}$

where $\left\| \cdot\right\|_{2}$ is the Euclidean norm and $\left| \cdot\right|$ is the set cardinality.

**Statistical analysis**

**Ablation studies.** For ablation, we fit linear mixed effects models for each dataset independently, with random intercepts to compare changes in both Dice scores and HD between our proposed CNN and each corresponding ablation (24). Each CNN was represented as a fixed effect with the unablated CNN serving as the reference category. With this approach, model coefficients solely reflect difference in subject-averaged Dice scores between each ablated CNN and the unablated CNN. P-values for the linear mixed effects model were derived as the significance of a two-tailed Wald z-test with a null hypothesis of zero-slope and zero-intercept model coefficients. We considered all p-values of less than 0.05 statistically significant. We chose a linear mixed effects model because the ablation analysis consisted of repeat (and thus dependent) measures of Dice scores and HDs with varying modifications to the same basic CNN architecture. The linear mixed effects model and corresponding p-values were calculated with the Python *statsmodels* library.

**Test-retest analysis.** For test-retest analysis, we chose the reliability metric as the ICC. ICC was calculated with a two-way ANOVA mixed effects model based on a single volume measurement for each WM bundle (25). For assessment of correlation between WM bundle volumes and ICCs, we utilized a coefficient of determination derived from linear least-squares regression (R^2^) and calculated the significance (p-value) based on a two-tailed Wald t-test for a zero-slope regression null hypothesis. ICCs were calculated with the Python *pingouin* library, and R^2^/Wald t-test p-values were calculated with the Python *scipy* library.

**Analysis of AD, PD, and TBI datasets.** We used a two-tailed Wilcoxon rank-sum test (26) to assess group-wise FA and volumetric alterations in all ROIs chosen for clinical analysis. One exception for group-wise statistical-significance testing was for PD subject groups from the PPMI dataset, which consisted of two scans from the same subject. Here we used a two-tailed Wilcoxon signed-rank test for evaluation of FA and volumetric alterations in ROIs between the baseline and 2YFU PD subject groups. All p-values were corrected for multiple-comparisons across all assessed ROIs (e.g., n=16 for brainstem WM bundles) using the Benjamini-Hochberg procedure for false discovery rate control (27). We chose to use nonparametric statistical models due to 1. variable sample-sizes, 2. to avoid normality and homoscedasticity assumptions, and 3. to increase robustness for outliers, which we have empirically found to be more dominant when analyzing small WM bundle ROIs. All raw p-values were calculated with the Python *scipy* library, and multiple-comparisons correction was performed with the Python *statsmodels* library. We tested the discriminatory power of group-wise classification in the AD, PD and TBI datasets with cross-validated LDA due to its simplicity, linearity, and minimal assumptions about the underlying distribution of the data causing classification performance to be a direct reflection of the input features (i.e., average FA and volume). Prior to classifier training, we standardized all feature vectors with Z-scoring (centering to a zero-mean and scaling to a unit-variance). To account for the limited subject numbers in our datasets, and to avoid overfitting, we used leave-one-out cross-validation during training of each classifier. Classifiers were optimized with the Limited-Memory Broyden-Fletcher-Goldfarb-Shanno (LBFGS) algorithm (28). For consistency, we also used the same feature standardization method LDA classification model for ROC analysis of brainstem masks (represented as a scalar feature resulting in a one-dimensional boundary problem). We generated ROC curves by varying decision boundary for each classifier, and we chose AUC as the ROC performance metric. LDA classifier setup, training, and ROC curve generation was performed with the Python *scikit-learn* library. The nonparametric significance model used to compare AUCs between two ROC curves was chosen to be the uncorrected two-tailed paired DeLong test (29). DeLong p-value calculation was performed in python with functions adapted from the Video Multi-Method Assessment Fusion (VMAF) library (30)

**Ablation Dice score summaries for linear mixed-effects models with random intercepts**

```
HCP Model Summary:
                 Mixed Linear Model Regression Results
=======================================================================
Model:                  MixedLM      Dependent Variable:      DiceScore
No. Observations:       75           Method:                  REML
No. Groups:             15           Scale:                   0.0003
Min. group size:        5            Log-Likelihood:          152.7088
Max. group size:        5            Converged:               Yes
Mean group size:        5.0
-----------------------------------------------------------------------
                             Coef.  Std.Err.   z    P>|z| [0.025 0.975]
-----------------------------------------------------------------------
Intercept                     0.698    0.011 64.156 0.000  0.677  0.719
CNN[T.Ablated_CRF CNN]       -0.005    0.007 -0.814 0.415 -0.018  0.008
CNN[T.Ablated_Attention CNN] -0.059    0.007 -8.968 0.000 -0.072 -0.046
CNN[T.PFM V1 CNN]            -0.028    0.007 -4.265 0.000 -0.041 -0.015
CNN[T.Ablated_PFM CNN]       -0.046    0.007 -6.945 0.000 -0.059 -0.033
Group Var                     0.001    0.035
=======================================================================
Ex Vivo Model Summary:
                 Mixed Linear Model Regression Results
========================================================================
Model:                   MixedLM      Dependent Variable:      DiceScore
No. Observations:        35           Method:                  REML
No. Groups:              7            Scale:                   0.0007
Min. group size:         5            Log-Likelihood:          52.7506
Max. group size:         5            Converged:               Yes
Mean group size:         5.0
------------------------------------------------------------------------
                             Coef.  Std.Err.    z    P>|z| [0.025 0.975]
------------------------------------------------------------------------
Intercept                     0.615    0.020  30.453 0.000  0.576  0.655
CNN[T.Ablated_CRF CNN]       -0.025    0.014  -1.764 0.078 -0.054  0.003
CNN[T.Ablated_Attention CNN] -0.138    0.014  -9.626 0.000 -0.167 -0.110
CNN[T.PFM V1 CNN]            -0.105    0.014  -7.280 0.000 -0.133 -0.077
CNN[T.Ablated_PFM CNN]       -0.148    0.014 -10.306 0.000 -0.176 -0.120
Group Var                     0.002    0.055
========================================================================
ADNI Model Summary:
                 Mixed Linear Model Regression Results
=======================================================================
Model:                  MixedLM      Dependent Variable:      DiceScore
No. Observations:       50           Method:                  REML
No. Groups:             10           Scale:                   0.0002
Min. group size:        5            Log-Likelihood:          106.5039
Max. group size:        5            Converged:               Yes
Mean group size:        5.0
-----------------------------------------------------------------------
                             Coef.  Std.Err.   z    P>|z| [0.025 0.975]
-----------------------------------------------------------------------
Intercept                     0.664    0.010 65.586 0.000  0.645  0.684
CNN[T.Ablated_CRF CNN]       -0.023    0.007 -3.466 0.001 -0.036 -0.010
CNN[T.Ablated_Attention CNN] -0.045    0.007 -6.834 0.000 -0.058 -0.032
CNN[T.PFM V1 CNN]            -0.062    0.007 -9.301 0.000 -0.075 -0.049
CNN[T.Ablated_PFM CNN]       -0.062    0.007 -9.326 0.000 -0.075 -0.049
Group Var                     0.001    0.030
=======================================================================
```

**Ablation Haussdorf distance summaries for linear mixed-effects models with random intercepts**

```
HCP Model Summary:
                 Mixed Linear Model Regression Results
=======================================================================
Model:                  MixedLM       Dependent Variable:       HDScore
No. Observations:       75            Method:                   REML
No. Groups:             15            Scale:                    0.0190
Min. group size:        5             Log-Likelihood:           16.9541
Max. group size:        5             Converged:                Yes
Mean group size:        5.0
-----------------------------------------------------------------------
                             Coef.  Std.Err.   z    P>|z| [0.025 0.975]
-----------------------------------------------------------------------
Intercept                     2.108    0.058 36.251 0.000  1.994  2.222
CNN[T.Ablated_CRF CNN]       -0.009    0.050 -0.182 0.856 -0.108  0.090
CNN[T.Ablated_Attention CNN]  0.081    0.050  1.599 0.110 -0.018  0.179
CNN[T.PFM V1 CNN]             0.058    0.050  1.147 0.251 -0.041  0.156
CNN[T.Ablated_PFM CNN]        0.103    0.050  2.053 0.040  0.005  0.202
Group Var                     0.032    0.109
=======================================================================
Ex Vivo Model Summary:
                 Mixed Linear Model Regression Results
=======================================================================
Model:                  MixedLM       Dependent Variable:       HDScore
No. Observations:       35            Method:                   REML
No. Groups:             7             Scale:                    0.0362
Min. group size:        5             Log-Likelihood:           -5.2527
Max. group size:        5             Converged:                Yes
Mean group size:        5.0
-----------------------------------------------------------------------
                             Coef.  Std.Err.   z    P>|z| [0.025 0.975]
-----------------------------------------------------------------------
Intercept                     2.322    0.131 17.708 0.000  2.065  2.580
CNN[T.Ablated_CRF CNN]       -0.051    0.102 -0.504 0.614 -0.250  0.148
CNN[T.Ablated_Attention CNN]  0.201    0.102  1.977 0.048  0.002  0.400
CNN[T.PFM V1 CNN]             0.413    0.102  4.061 0.000  0.214  0.612
CNN[T.Ablated_PFM CNN]        0.590    0.102  5.806 0.000  0.391  0.789
Group Var                     0.084    0.310
=======================================================================
ADNI Model Summary:
                Mixed Linear Model Regression Results
======================================================================
Model:                   MixedLM      Dependent Variable:      HDScore
No. Observations:        50           Method:                  REML
No. Groups:              10           Scale:                   0.0125
Min. group size:         5            Log-Likelihood:          13.0331
Max. group size:         5            Converged:               Yes
Mean group size:         5.0
----------------------------------------------------------------------
                             Coef. Std.Err.   z    P>|z| [0.025 0.975]
----------------------------------------------------------------------
Intercept                    2.336    0.099 23.651 0.000  2.142  2.529
CNN[T.Ablated_CRF CNN]       0.031    0.050  0.614 0.539 -0.067  0.129
CNN[T.Ablated_Attention CNN] 0.003    0.050  0.050 0.960 -0.095  0.100
CNN[T.PFM V1 CNN]            0.082    0.050  1.638 0.101 -0.016  0.180
CNN[T.Ablated_PFM CNN]       0.136    0.050  2.730 0.006  0.038  0.234
Group Var                    0.085    0.413
```

======================================================================

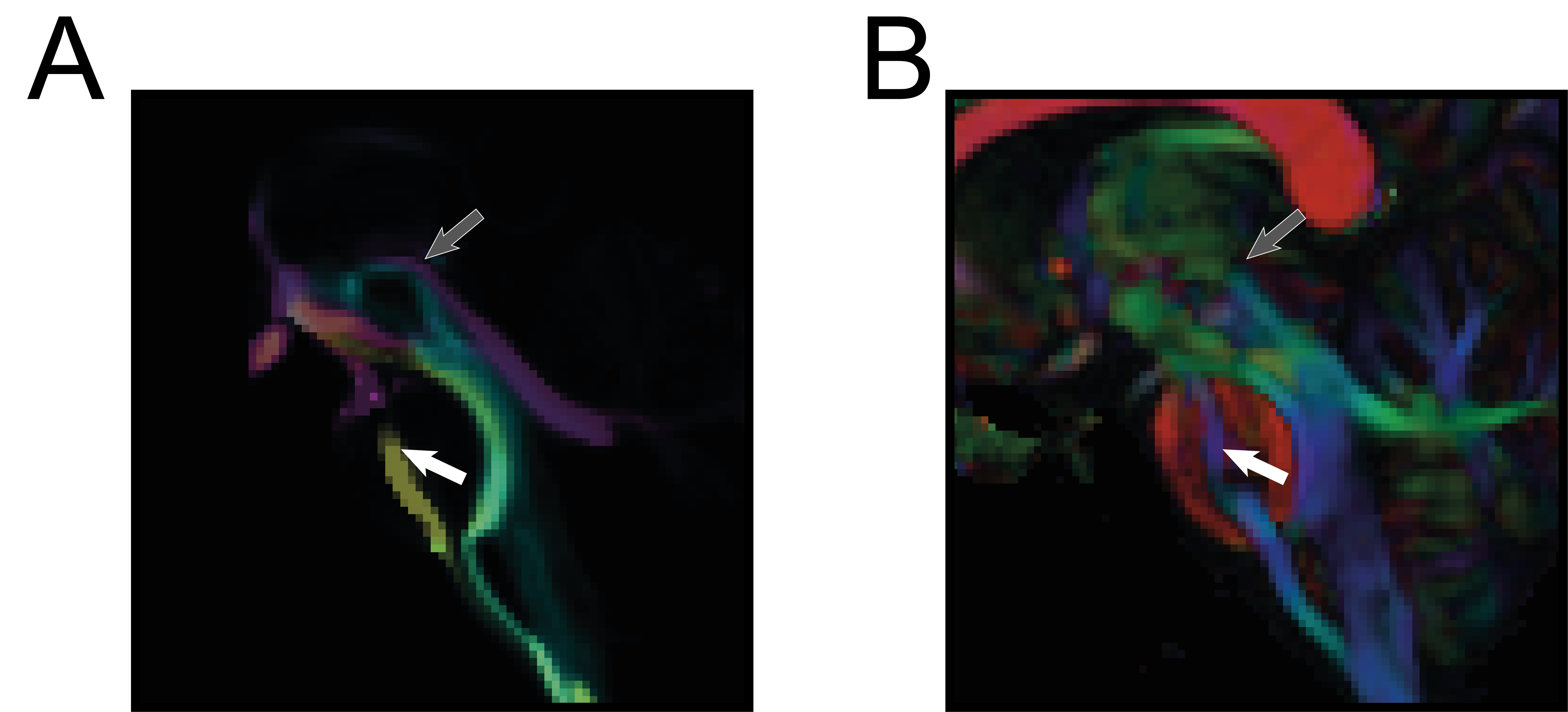

**Fig. S1**. Bundle coherence with streamline intensity mapping. A sagittal section of a PFM channel is shown in (A), with a corresponding color FA map of the same section displayed in (B). Large fiber bundles such as the corticospinal pathways (white arrow) are easily-discernable with inspection of either the color FA map or the PFM channel. However, bundles in the midbrain with smaller cross-sectional areas and more tortuosity (grey arrow) require a tractographic approach to generate reliable contrast, which is not captured solely by inspection of local diffusion directionality.

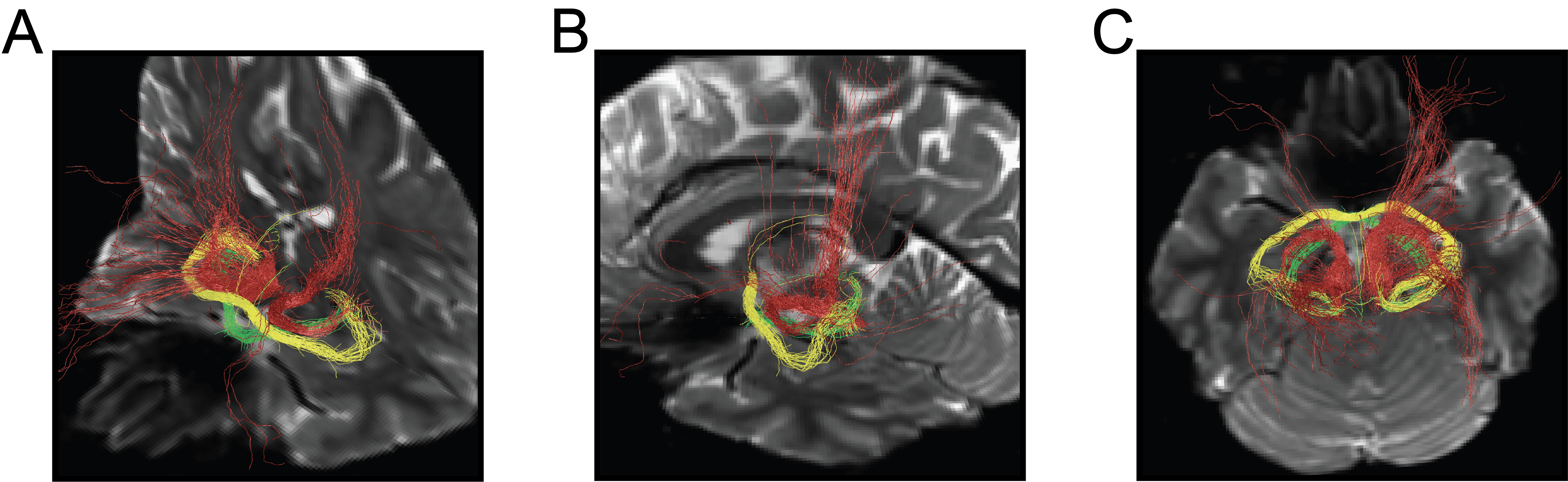

**Fig. S2**. Deterministic tractography of the lateral forebrain bundle in the brainstem. Oblique (A), mid-sagittal (B) and axial (C) views of deterministic streamlines generated with *MRtrix* software (31) display the architecture of the LFB (brown), as confirmed with previous descriptions in humans (3). LFB streamlines are constrained to streamlines that pass through all regularly-spaced axial cross-sections of the BSB LFB segmentation. Streamlines generated from anterior commissure (yellow) and optic tract (green) regions of interest confirm that the LFB reconstruction forms unique and distinct extra-brainstem projections.

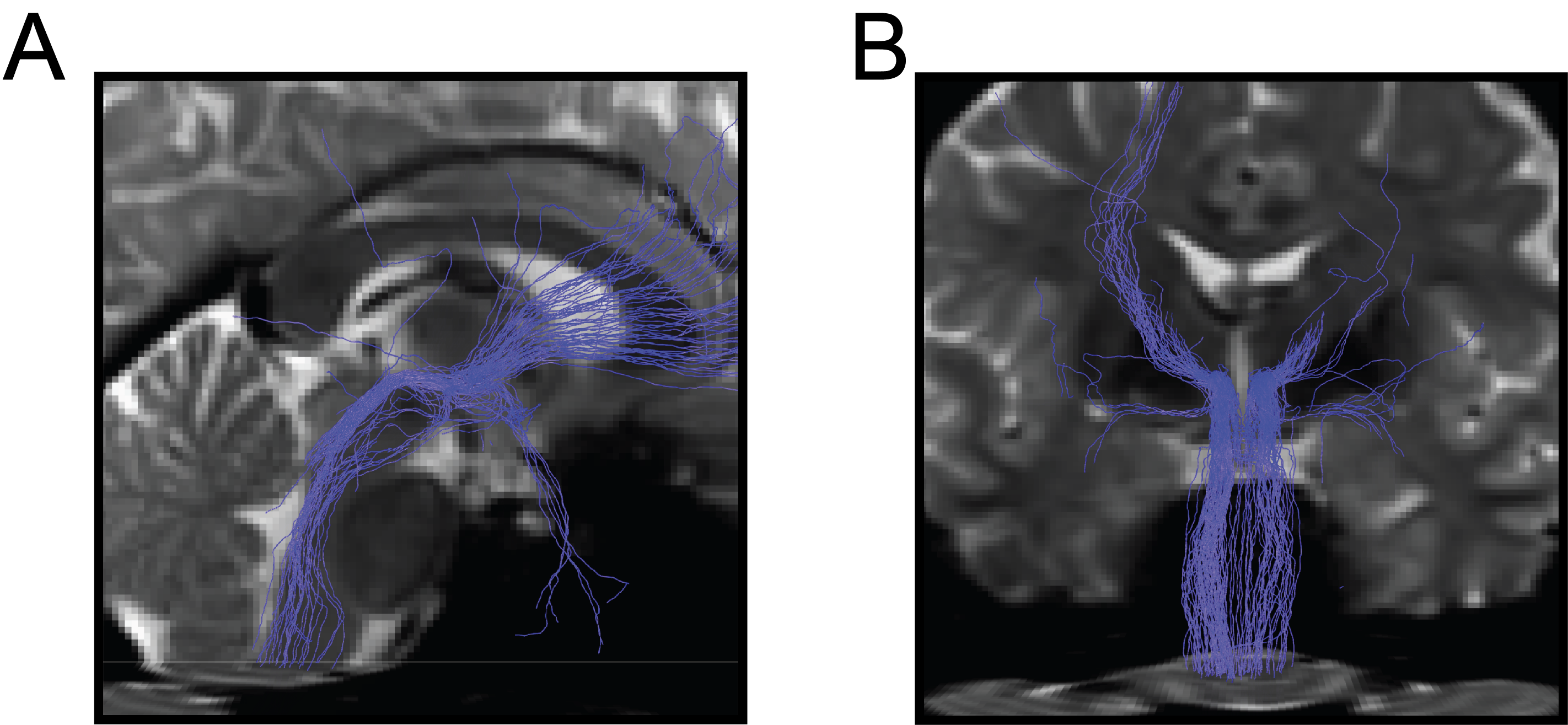

Fig. S3. Deterministic tractography of the mesencephalic homeostatic bundle in the brainstem. Representative mid-sagittal (A) and coronal (B) views of the deterministic streamlines passing through and emanating from the BSB MHB segmentation. The MHB architecture was confirmed with a prior study of MHB anatomy in humans (3).

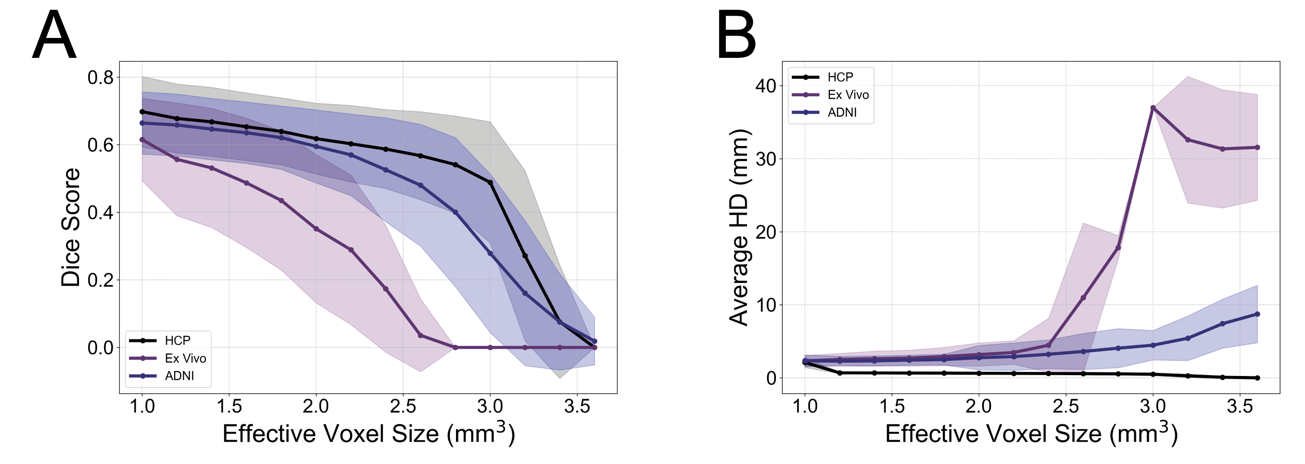

**Fig. S4**. Effect of spatial resampling on BSBT accuracy. We tested the robustness of BSBT segmentation to differing spatial resolution. To mimic resolution changes, we apply gaussian smoothing with a standard deviation of $\sigma=r_{target}/\left( r_{native}\cdot\sqrt{2\cdot log(2)} \right)$, where $r_{target}$ and $r_{native}$ are the target and native dMRI resolutions respectively. Dice scores and aHD averaged over all brainstem WMB are shown in panels (A) and (B) respectively. For each resampling step, missing segmentation labels (i.e., where a label volume is zero), were given a Dice score of 0 and excluded from average HD calculation, as a zero-label would lead to an infinite HD.

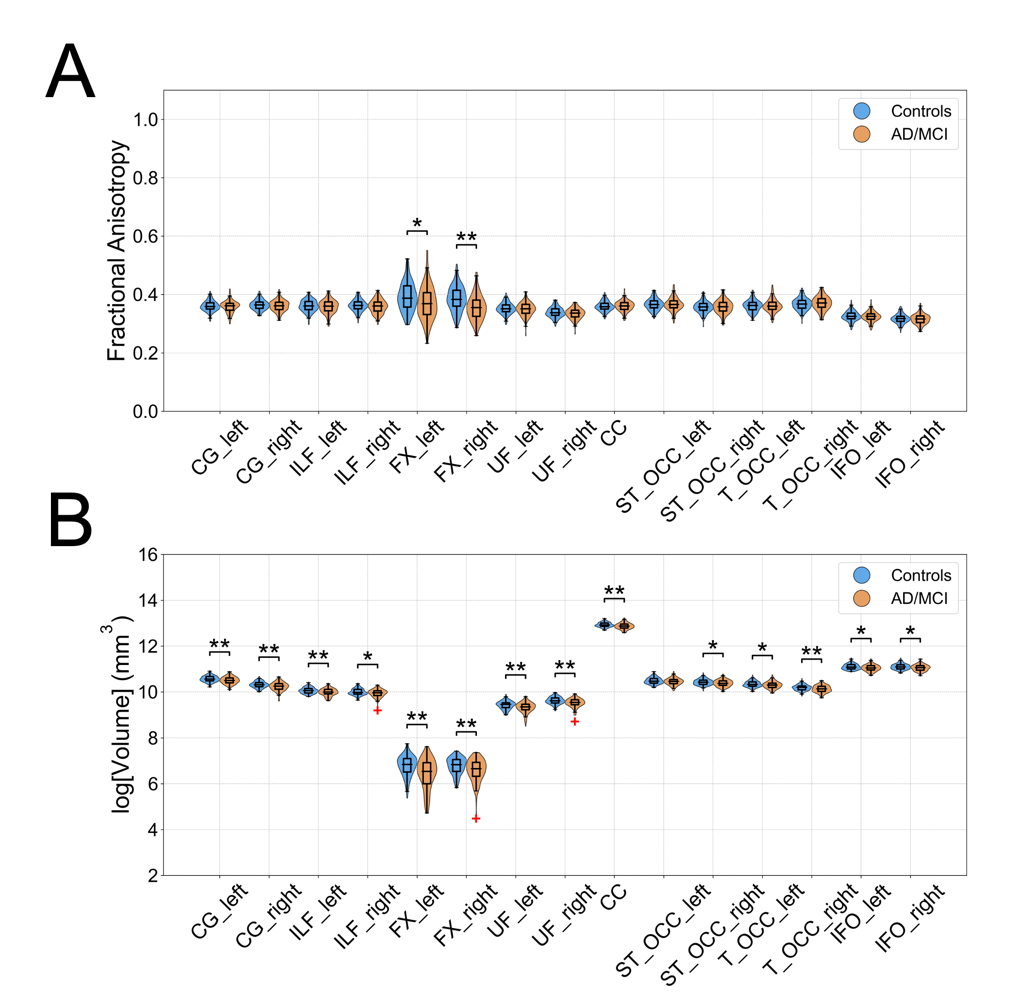

**Fig. S5**. Supra-tentorial white matter alterations in Alzheimer's disease/mild cognitive impairment. Violin plots for average FA (A) and log-volume (B) for the *TractSeg*-derived supra-tentorial WM bundles in healthy controls (blue) and AD/MCI patients (orange) from the ADNI3 dataset. Significance bars indicate FDR-corrected two-tailed Wilcoxon rank-rum p value of <0.05 (*) or <0.01 (**). CG: Cingulum, ILF: Inferior Longitudinal Fasciculus, FX: Fornix, UF: Uncinate fasciculus, CC: Corpus Callosum, ST_OCC: Striato-Occipital tract, T_OCC: Thalamo-Occipital tract, IFO: Inferior Occipito-Frontal fasciculus.

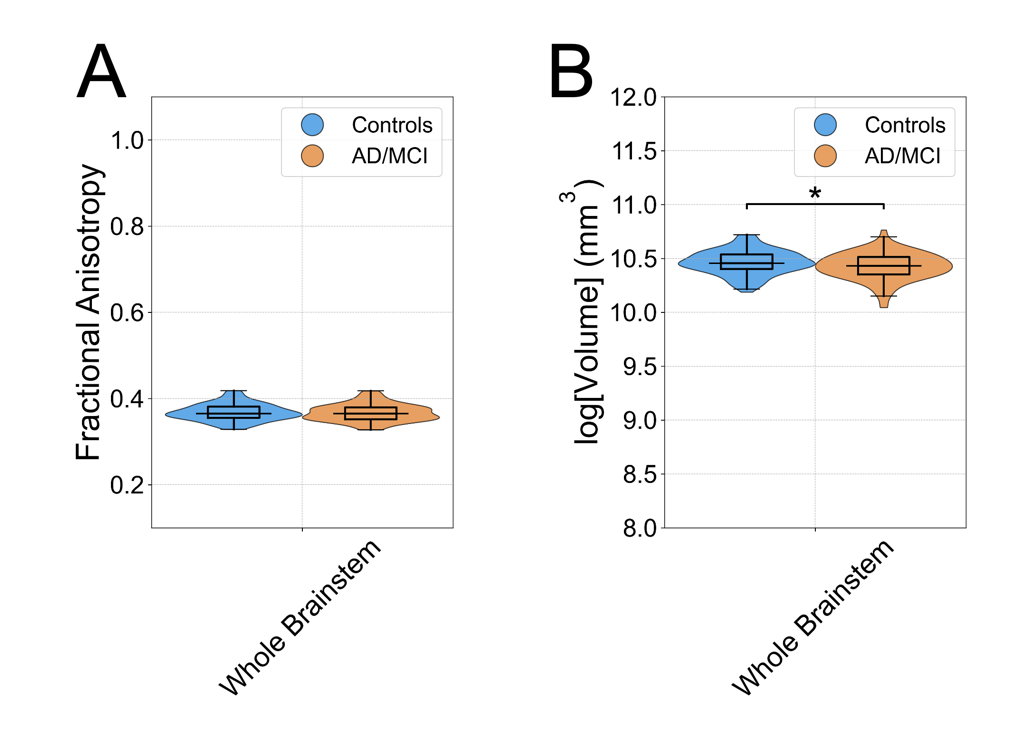

**Fig. S6**. Whole-brainstem alterations in Alzheimer's disease/mild cognitive impairment. Violin plots for average FA (A) and log-volume (B) for the whole brainstem mask in healthy controls (blue) and AD/MCI patients (orange) from the ADNI3 dataset. Significance bars indicate an FDR-corrected two-tailed Wilcoxon rank-rum p value of <0.05 (*) or <0.01 (**).

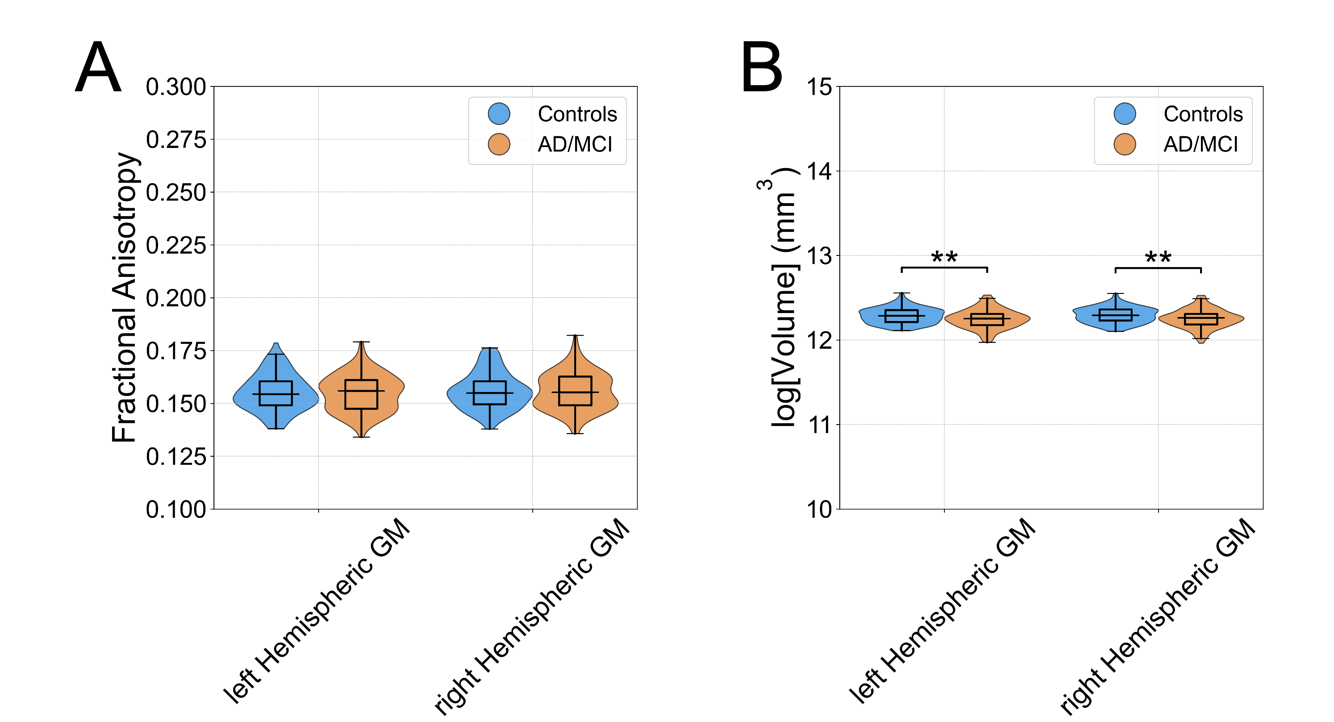

**Fig. S7**. Hemispheric grey matter alterations in Alzheimer's disease/mild cognitive impairment. Violin plots for average FA (A) and log-volume (B) for the hemispheric grey matter masks in healthy controls (blue) and AD/MCI patients (orange) from the ADNI3 dataset. Significance bars indicate an FDR-corrected two-tailed Wilcoxon rank-rum p value of <0.05 (*) or <0.01 (**).

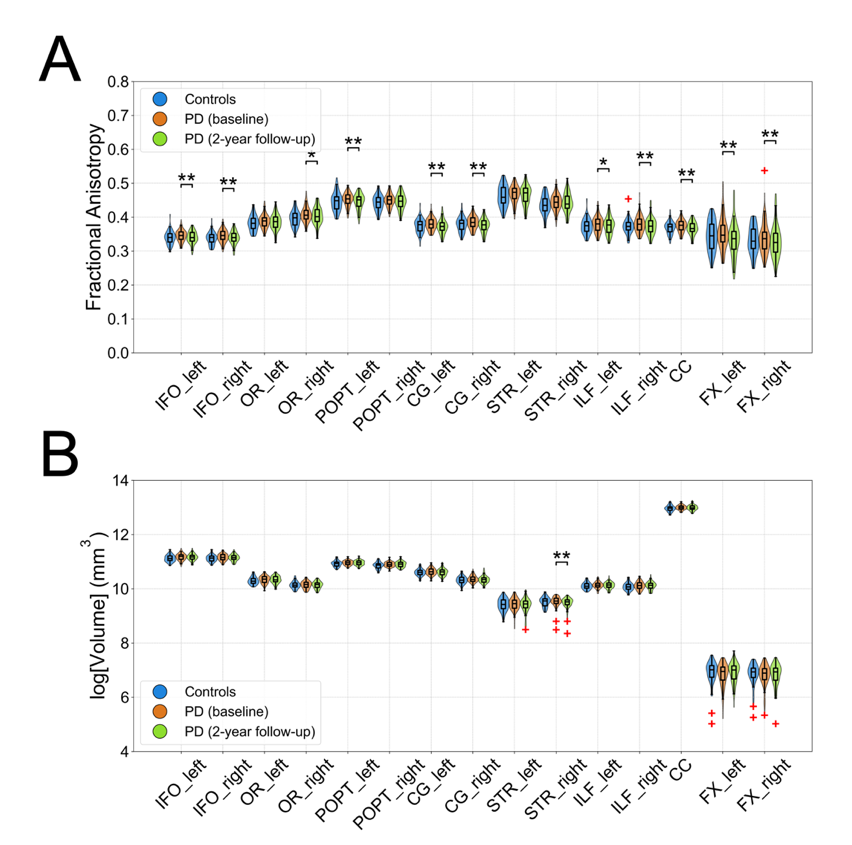

**Fig. S8**. Supra-tentorial white matter alterations in Parkinson's disease. Violin plots of average FA (A) and log-volume (B) for *TractSeg*-derived supra-tentorial WM bundles in control subjects and PD patients scanned at time of diagnosis and at a two-year follow-up session. Significance bars indicate an FDR-corrected two-tailed Wilcoxon rank-rum p value for comparison with controls and an FDR-corrected two-taed Wilcoxon signed-rank p value for comparison of PD baseline with two-year follow-up scans of <0.05 (*) or <0.01 (**). IFO: Inferior Occipito-Frontal Fasciculus. OR: Optic Radiation. POPT: Parieto-Occipital Pontine tracts. CG: Cingulum. STR: Superior Thalamic Radiation. ILF: Inferior Longitudinal Fasciculus. CC: Corpus Callosum. FX: Fornix.

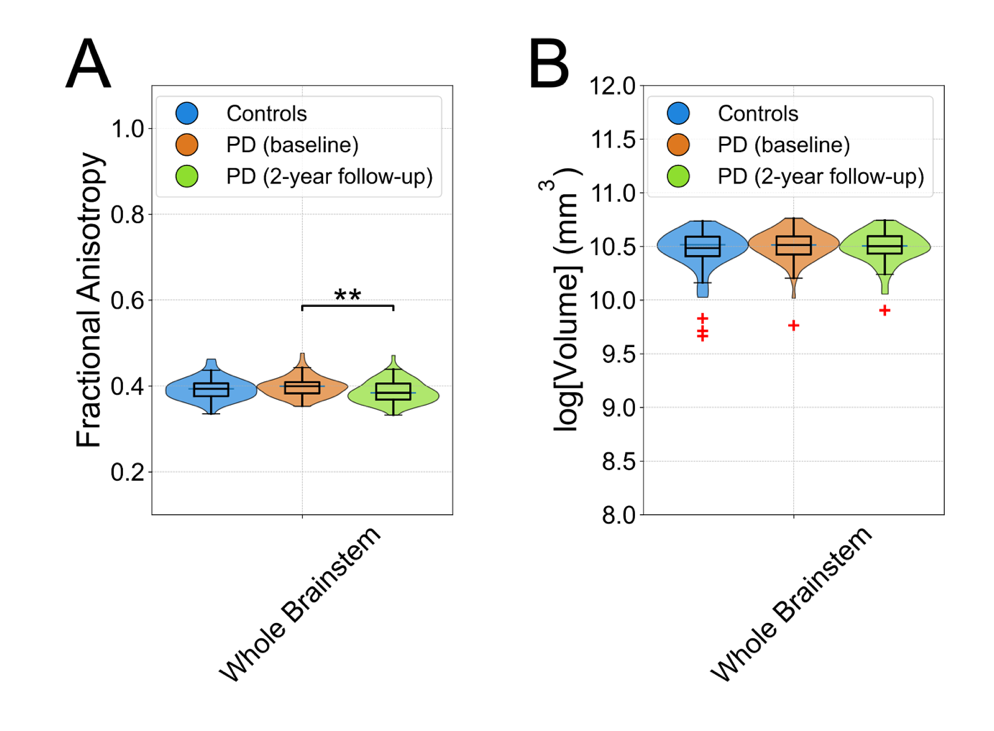

**Fig. S9**. Whole-brainstem alterations in Parkinson's disease. Violin plots of average FA (A) and log-volume (B) for the whole brainstem in control subjects and PD patients scanned at time of diagnosis and at a two-year follow-up *session*. Significance bars indicate an FDR-corrected two-tailed Wilcoxon rank-rum p value for comparison with controls or a Wilcoxon signed-rank p value for comparison of PD baseline with two-year follow-up scans of <0.05 (*) or <0.01 (**).

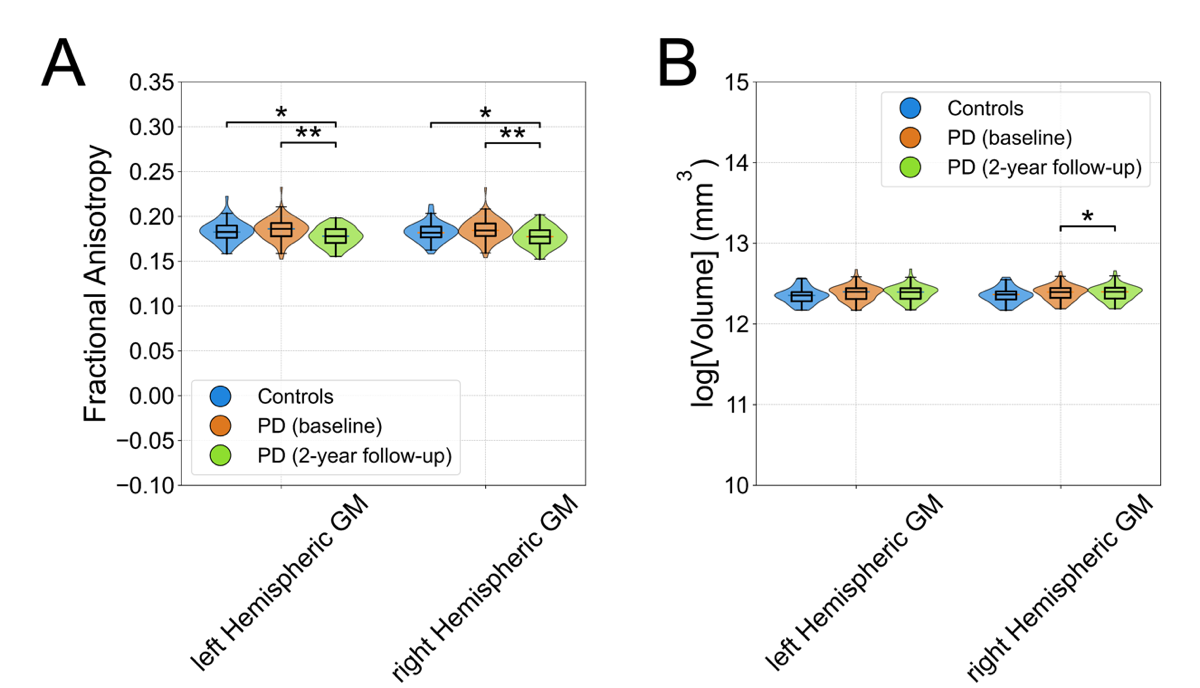

**Fig. S10**. Hemispheric grey matter alterations in Parkinson's disease. Violin plots of average FA (A) and log-volume (B) for the hemispheric GM masks in control subjects and PD patients scanned at time of diagnosis and at a two-year follow-up session. Significance bars indicate an FDR-corrected two-tailed Wilcoxon rank-rum p value for comparison with controls or an FDR-corrected two-tailed Wilcoxon signed-rank p value for comparison of PD baseline with two-year follow-up scans of <0.05 (*) or <0.01 (**).

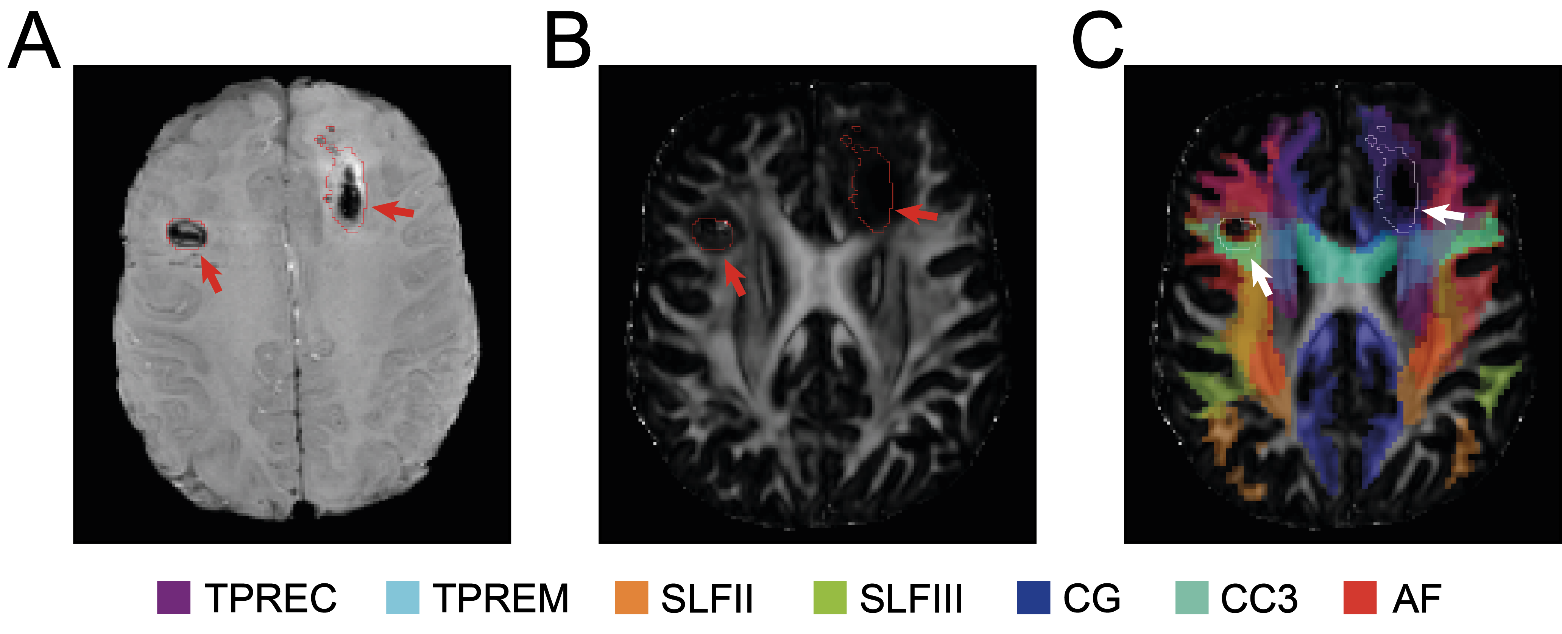

**Fig. S11**. Mapping of manually annotated TBI lesions with white matter segmentations. Hypointense hemorrhagic lesions were manually annotated in the SWI scans of each TBI patient (A) that were registered with an affine transformation model into dMRI space (B). (C) Supra-tentorial white matter bundles segmented with *TractSeg* were then overlayed with the lesion regions, and white matter bundles with the most lesion overlap were chosen for LR classification comparison with BSBT WM bundles.

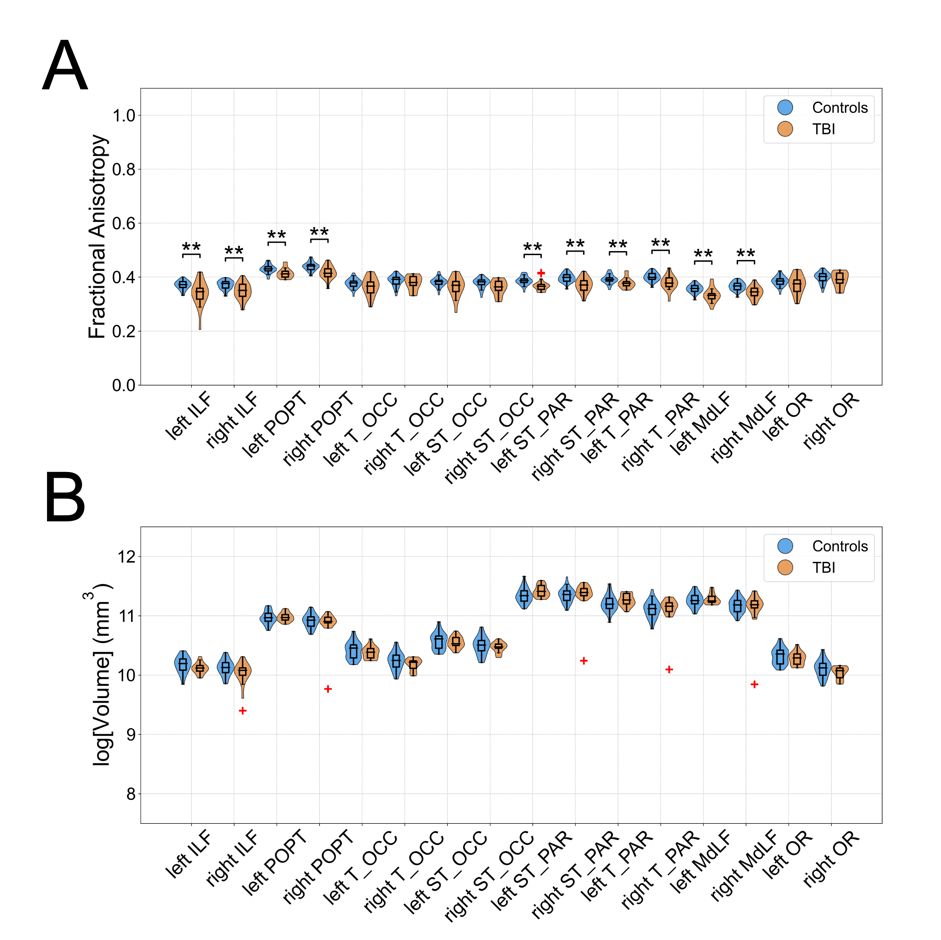

**Fig. S12**. Supra-tentorial white matter alterations in traumatic brain injury. Violin plots for average FA (A) and log-volume (B) for the *TractSeg*-derived supra-tentorial WM bundles with the greatest degree of SWI lesion overlap in healthy controls (blue) and acute TBI patients (orange) from the TBI dataset. Significance bars indicate an FDR-corrected two-tailed Wilcoxon rank-rum p value of <0.05 (*) or <0.01 (**). CG: Cingulum, ILF: Inferior Longitudinal Fasciculus, POPT: Parieto-Occipital Pontine tract, T_OCC: Thalamo-Occipital tract, ST_OCC: Striato-Occipital tract, ST_PAR: Striato-Parietal tract, T_PAR: Thalamo-Parietal tract, MdLF: Middle Longitudinal fasciculus, OR: Optic Radiation.

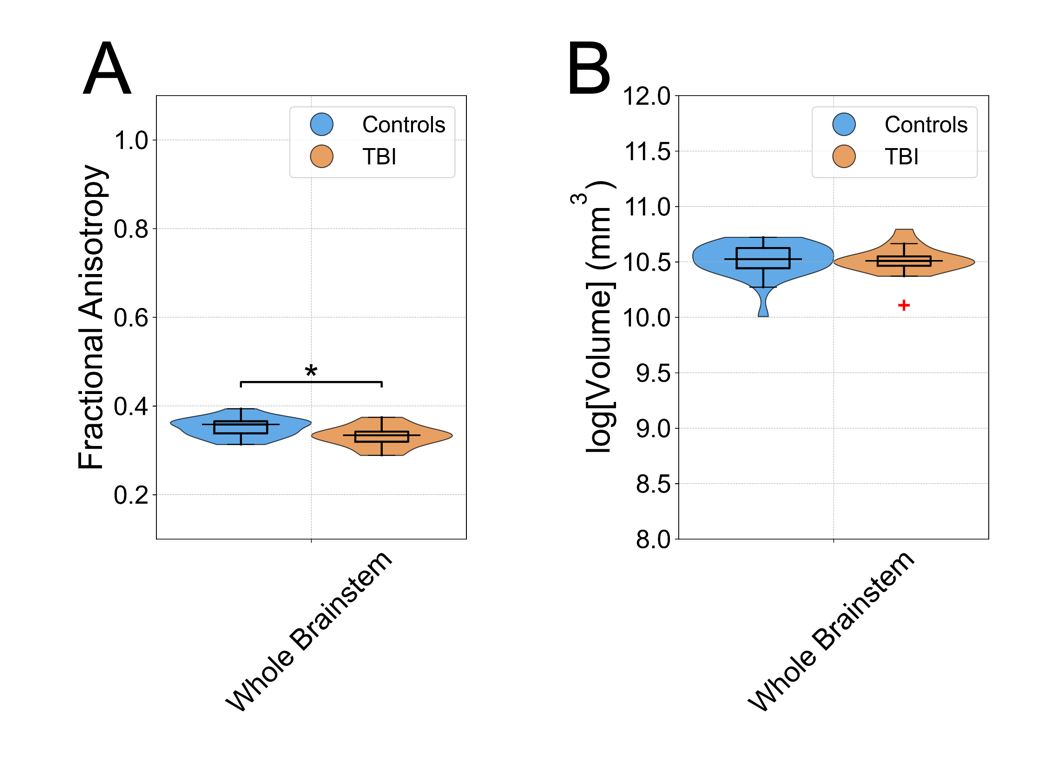

**Fig. S13**. Whole-brainstem alterations in traumatic brain injury. Violin plots for average FA (A) and log-volume (B) for the whole brainstem in healthy controls (blue) and TBI patients (orange) from the TBI dataset. Significance bars indicate an FDR-corrected two-tailed Wilcoxon rank-rum p value of <0.05 (*) or <0.01 (**).

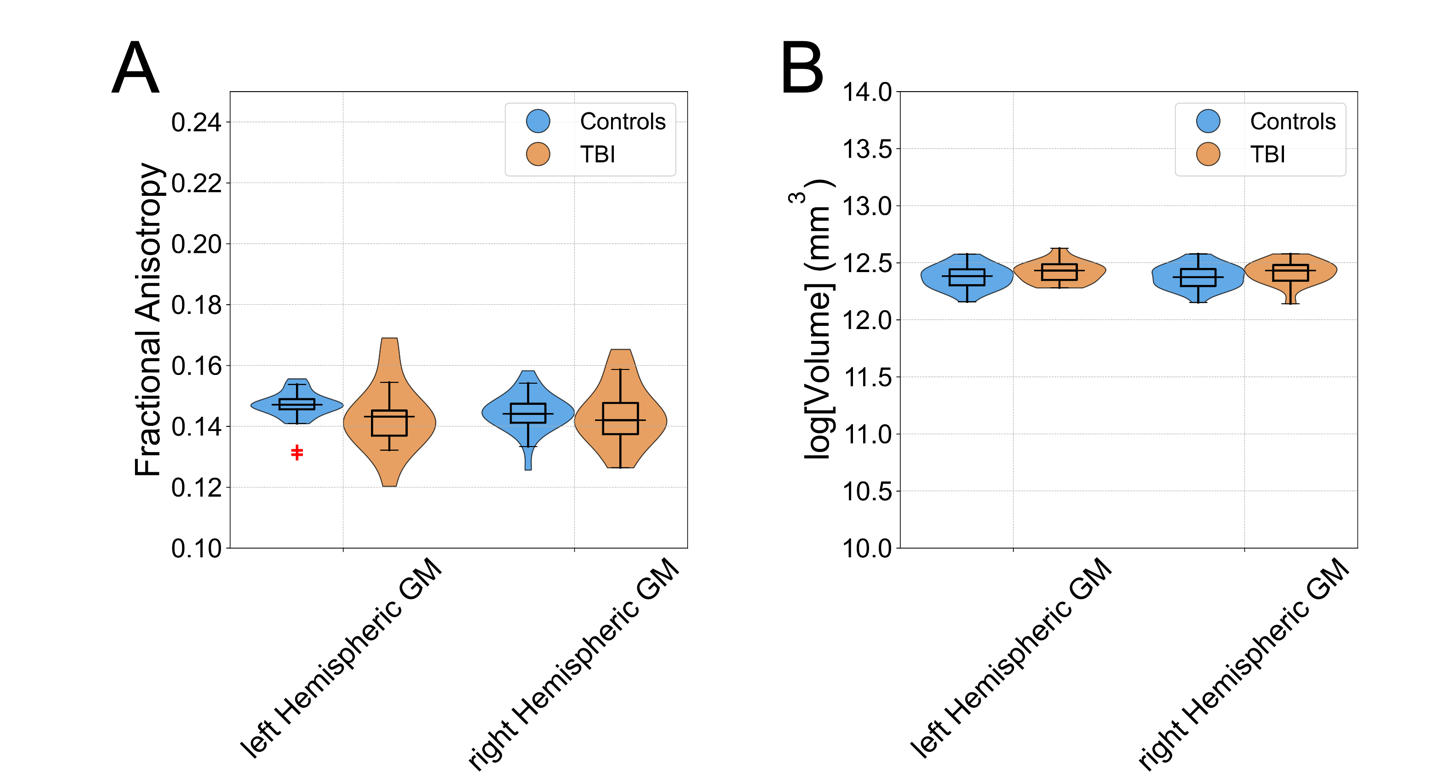

**Fig. S14**. Hemispheric grey matter alterations in traumatic brain injury. Violin plots for average along-tract MD for the hemispheric GM masks in healthy controls (blue) and TBI patients (orange) from the TBI dataset. Significance bars indicate a Wilcoxon rank-rum p value of <0.05 (*) or <0.01 (**).

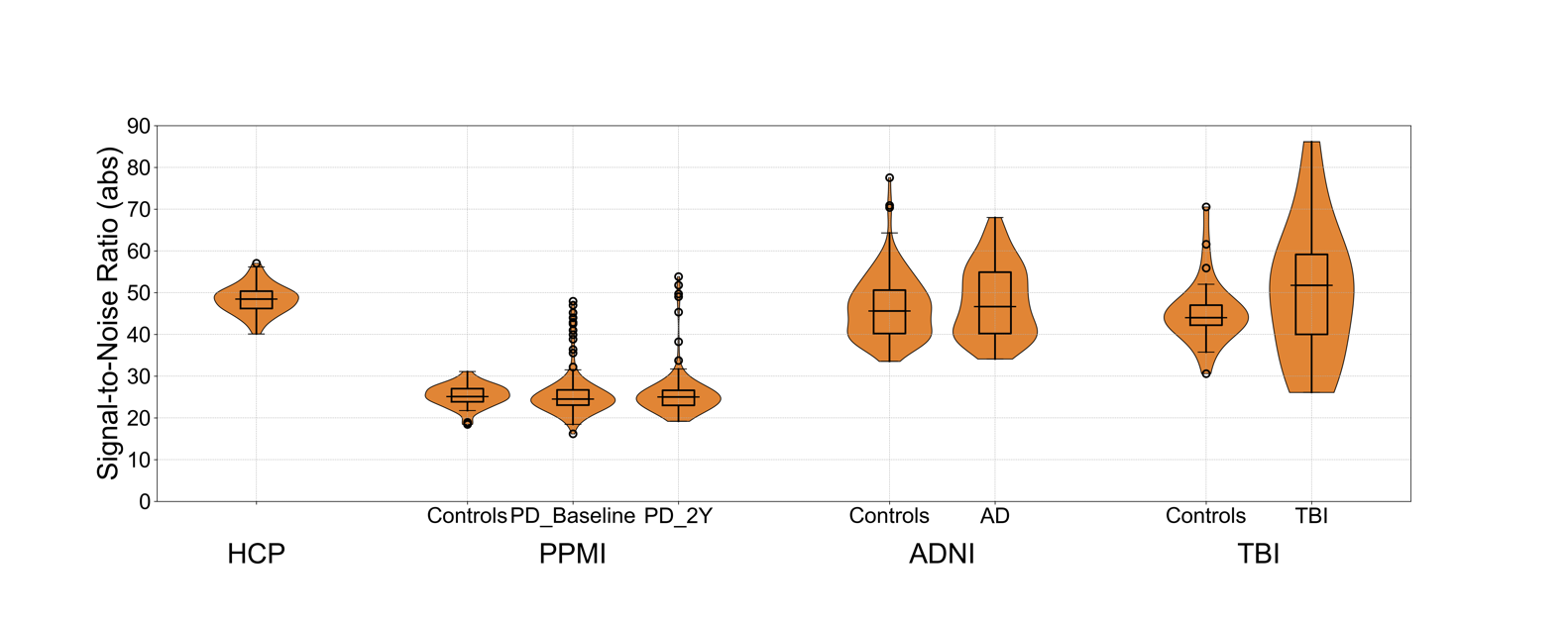

**Fig. S15**. Signal-to-noise ratios of clinical dMRI datasets. Violin plots with overlayed box plots indicating the median, 25th percentile, and 75th percentile values of signal-to-noise ratios calculated for each subject from the HCP, PPMI, ADNI and TBI datasets. The signal-to-noise ratio for each dMRI is calculated (in absolute space) as the ratio of the voxel-wise average intensity of the mean low-b to the voxel-wise average of the estimated noise map of the dMRI volume. Both voxel-wise averages are restricted to a brain mask. The estimated noise map is calculated via Marchenko-Pastur PCA denoising using the MRtrix *dwidenoise* command (cite). PD 2Y: PPMI PD 2-year follow-up scans. All dMRI data, including data with failed WM segmentations, in included in the above plots.

**Overview of *ex vivo* brain specimens used for histologically guided annotations, validation and ablation analysis**

| **ID** | **Age** | **Sex** | **Past Medical History** | **Cause of Death** | **Post-Mortem Fixation Interval (hours)** | **Fixation-to-Imaging Interval (months)** | **Fixative Used** | **Histology Included?** |
| --- | --- | --- | --- | --- | --- | --- | --- | --- |
| S1 | 81-85 | M | HLD, chronic respiratory deficiency | Acute respiratory failure | 24 | 50 | 5% formalin | no |
| S2 | 91-95 | M | HTN, CKD | No precise cause of death | 24 | 26 | 5% formalin | no |
| S4 | 91-95 | M | Lymphoma, T2DM, Prostate adenocarcinoma, DVT, Macrocytic anemia | Cardiac arrest with atrial fibrillation | 28 | 46 | 5% formalin | no |
| S4 | 61-65 | F | GI cancer, DVT | Septic shock | <24 | 24 | 10% formalin | yes |
| S5 | 61-65 | F | HTN, metastatic ovarian carcinoma | Septic shock | 72 | 20 | 10% formalin | yes |
| S6 | 46-50 | M | DVT, Raynaud’s phenomena, Leukemia | DIC due to Hemophagocytic lymphohistiocytosis | <24 | 93 | 10% formalin | no |
| S7 | 56-60 | F | Ulcerative colitis, Breast cancer | PE and development of DAD in setting of widely metastatic breast cancer | <24 | 92 | 10% formalin | no |

**Table S1.** HTN: Hypertension. CKD: Chronic Kidney Disease. GI: Gastrointestinal. DVT: Deep Vein Thrombosis. HLD: Hyperlipidemia. T2DM: Type 2 Diabetes Mellitus. PE: Pulmonary Embolism. DIC: Disseminated Intravascular Coagulation. DAD: Diffuse Alveolar Damage.

**Overview of traumatic brain injury patients used for clinical analysis**

| **ID** | **Age** | **Sex** | **Mechanism of injury** | **Days in Coma** | **Days Until Command-Following** | **Day of MRI** | **LOC at MRI** | **GCS-T at MRI** | **CRS-R-T at MRI** | **DRS 6 month** | **GOS-E 6 month** |
| --- | --- | --- | --- | --- | --- | --- | --- | --- | --- | --- | --- |
| P1 | 26-30 | M | MVA | 1 | 1 | 16 | PTCS | 15 | 23 | 7 | 3 |
| P2 | 21-25 | M | Ped vs car | 1 | 6 | 1 | MCS- | 7 | 4 | 0 | 7 |
| P3 | 16-20 | F | MVA | 5 | 10 | 3 | Coma | 5 | 1 | 0 | 7 |
| P4 | 16-20 | M | Fall | 1 | 2 | 17 | PTCS | 14 | 23 | 0 | 7 |
| P5 | 31-35 | M | Fall | 6 | 26 | 15 | VS | 7 | 3 | 8 | 3 |
| P6 | 26-30 | F | MVA | 2 | 9 | 7 | VS | 9 | 6 | 5 | 3 |
| P7 | 41-45 | M | MVA | 1 | 6 | 13 | MCS+ | 13 | 18 | 4 | 5 |
| P8 | 31-35 | M | Fall | 1 | 7 | 8 | PTCS | 11 | 20 | 5 | 3 |
| P9 | 31-35 | M | Ped vs car | 1 | 8 | 11 | MCS+ | 10 | 9 | 5 | 4 |
| P10 | 21-25 | M | Assault | 1 | 7 | 12 | MCS- | 10 | 10 | 3 | 5 |
| P11 | 21-25 | F | Ped vs car | 1 | 9 | 14 | PTCS | 14 | 22 | 0 | 7 |
| P12 | 26-30 | F | Fall | 13 | ** | 8 | Coma | 5 | 1 | 30 | 1 |
| P13 | 16-20 | M | Fall | 1 | 2 | 4 | MCS+ | 10 | 12 | 0 | 7 |
| P14 | 51-55 | M | Ped vs car | 8 | ** | 8 | VS | 6 | 3 | 30 | 1 |
| P15 | 26-30 | M | Ped vs car | 3 | 8 | 7 | MCS- | 7 | 3 | 3 | 5 |
| P16 | 31-35 | M | Fall | 3 | 3 | 3 | MCS+ | 10 | 12 | 2 | 5 |
| P17 | 26-30 | F | Ped vs car | 4 | ** | 12 | VS | 8 | 3 | 30 | 1 |
| P18 | 26-30 | M | Fall | 1 | 0–2* | 28 | PTCS | 11 | 18 | 18 | 3 |

**Table S2.** Further information on the previously-published trial parameters can be found at Edlow et. al. 2017 (*49*). MVA: Motor-Vehicle Accident. Ped vs car: pedestrian hit by car. MCS-: Minimally-Conscious State without language function. MCS+: Minimally-Conscious State with evidence of language function. VS: Vegetative State. PTCS: Post-Traumatic Confusional State. LOC: level of consciousness. GCS-T: total Glasgow Coma Scale. GOS-E: Glasgow Coma Scale-Extended. CRS-R-T: Coma Recovery Scale-Revised-Total. *No exact date given by treating clinician(s). **Patient died after withdrawal of life-sustaining therapy and before command-following could be assessed. DRS: Disability Rating Scale. Inclusion criteria for the study were: (1) between 18-65 years of age (2) presenting with a Glasgow Coma Scale Index of <8 and (3) no eye opening for at least 24h following injury. Exclusion criteria for the study were: (1) any history of TBI or other neurological disorders (2) life expectancy of <6 months as determined by a treating clinician (3) presence of any metal object in the cranium that could cause MRI artefact and (4) Lack of English fluency.

**List of TractSeg-derived white matter bundles and abbreviations**

| **Tract Name** | **Abbreviation** |
| --- | --- |
| Arcuate fasciculus | AF |
| Anterior Thalamic Radiation | ATR |
| Anterior Commissure | CA |
| Cingulum | CG |
| Cortico-Spinal tracts | CST |
| Middle Longitudinal fasciculus | MdLF |
| Fronto-Pontine tract | FPT |
| Fornix | FX |
| Inferior Cerebellar Peduncle | ICP |
| Inferior Occipito-Frontal fasciculus | IFO |
| Inferior Longitudinal Fasciculus | ILF |
| Middle Cerebellar Peduncle | MCP |
| Optic Radiation | OR |
| Parieto-Occipital Pontine tract | POPT |
| (TractSeg-derived) Superior Cerebellar Peduncle | SCPt |
| Superior Longitudinal Fasciculus subdivisions I-III | SLFI-III |
| Superior Thalamic Radiation | STR |
| Uncinate Fasciculus | UF |
| Corpus Callosum | CC |
| Thalamo-Prefrontal tract | TPREF |
| Thalamo-Premotor tract | TPREM |
| Thalamo-Precentral tract | TPREC |
| Thalamo-postcentral tract | TPOSTC |
| Thalamo-Parietal tract | TPAR |
| Thalamo-Occipital tract | TOCC |
| Striato-Fronto-Orbital tract | STFO |
| Striato-Prefrontal tract | STPREF |
| Striato-Premotor tract | STPREM |
| Striato-Precentral tract | STPREC |
| Striato-Postcentral tract | STPOSTC |
| Striato-Parietal tract | STPAR |
| Striato-Occipital tract | STOCC |

**Table S3.** All abbreviations for *TractSeg* WM bundles can be found in the associated publication (32) and <https://github.com/MIC-DKFZ/TractSeg>. Abbreviations for the Middle Longitudinal fasciculus (MdLF) and Superior Cerebellar Peduncle (SCPt) were modified from their original descriptions to avoid naming ambiguity with the BSBT-derived MLF and SCP.

Dataset S1 (separate file). Excel file with all subject-specific information (including but not limited to age, sex, disease status, and inclusion/exclusion from processing) from the HCP, ADNI, TBI, and *ex vivo* brain specimen datasets.
